# Benefits and risks of zinc for adults during covid-19: rapid systematic review and meta-analysis of randomised controlled trials

**DOI:** 10.1101/2020.11.02.20220038

**Authors:** Jennifer Hunter, Susan Arentz, Joshua Goldenberg, Guoyan Yang, Jennifer Beardsley, Stephen P Myers, Dominik Mertz, Stephen Leeder

## Abstract

**Objective:** To evaluate the benefits and risks of any type of zinc intervention to prevent or treat SARS-CoV-2.

**Design:** A living, systematic review and meta-analysis, incorporating rapid review methods.

**Data sources:** 17 English and Chinese databases and clinical trial registries were searched in April/May 2020, with additional covid-19 focused searches in June and August 2020.

**Eligibility criteria and analysis:** Randomized control trials (RCTs) published in any language comparing zinc to a control to prevent or treat SARS-CoV-2. Other viral respiratory tract infections (RTIs) were included, but the certainty of evidence downgraded twice for indirectness. Screening, data extraction, risk of bias appraisal (RoB-2 tool) and verification was performed by calibrated, single reviewers. RCTs with adult populations were prioritised for analysis.

**Results:** 123 RCTs were identified. None were specific to SARS-CoV-2 nor other coronaviruses. 28 RCTs evaluated oral (15-45mg daily), sublingual (45-300mg daily), or topical nasal (0.09-2.6 mg daily) zinc to prevent or treat nonspecific viral RTIs in 3,597 adults without zinc deficiency. Compared to placebo, zinc prevented 5 mild to moderate RTIs per 100 person-months, including in older adults (95% confidence interval 1 to 9) (number needed to treat (NTT)=20). There was no significant difference in the rates of non-serious adverse events (AE). For RTI treatment, a clinically important reduction in peak symptom severity scores was found for zinc compared to placebo (mean difference 1.2 points, 0.7 to 1.7), but not average daily symptom severity (standardised mean difference 0.2, 0.1 to 0.4). 19 fewer per 100 adults were at risk of remaining symptomatic over the first 7 days (2 to 38, NNT=5) and the mean duration of symptoms was 2 days shorter (0.2 to 3.5), however, there was substantial heterogeneity (I^2^ = 82% and 97%). 14 more per 100 experienced a non-serious AE (4 to 16, NNT=7) such as nausea, or mouth or nasal irritation. No differences in illness duration nor AE were found when zinc was compared to active controls. No serious AE, including copper deficiency, were reported by any RCT. Quality of life outcomes were not assessed. Confidence in these findings for SARS-CoV-2 is very low due to serious indirectness and some concerns about bias for most outcomes.

**Conclusions:** Zinc is a potential therapeutic candidate for preventing and treating SARS-CoV-2, including older adults and adults without zinc deficiency (very low certainty). Zinc may also help to prevent other viral RTIs during the pandemic (moderate certainty) and reduce the severity and duration of symptoms (very low certainty). The pending results from seven RCTs evaluating zinc for SARS-CoV-2 will be tracked.

**Systematic review registration:** PROSPERO CRD42020182044

## BACKGROUND

In response to the global covid-19 pandemic, the Cochrane Collaboration developed a list of review priority questions^1^ and resources for conducting high quality rapid reviews^2^. Available antiviral, anti-inflammatory and anticoagulant pharmaceuticals are being evaluated^3^. Other interventions also being investigated include host-directed therapies and nutritional interventions. ^4^Zinc is one such intervention. By May 2020, 19 clinical trials evaluating zinc for the prevention or treatment of SARS-CoV-2 infections, either as a stand-alone therapy or combined with other pharmaceuticals or nutraceuticals, had been registered on at least one international clinical trial registry^5^.

Many people are not waiting for definitive evidence. Both high and low income countries have reported increased zinc supplement use and sales related to the covid-19 pandemic,^6 7^ including self-prescribing of prophylactic zinc by healthcare workers^8^ and some clinicians and hospitals using zinc to treat SARS-CoV-2 infections.^9-19^

Findings for zinc from five retrospective observational studies and a case series have reported mixed results. In India, no added protection against SARS-CoV-2 infection was found for healthcare workers who used various prophylactic nutritional interventions including zinc.^8^ In the United States, reduced in-hospital mortality was reported when 50mg to 100mg of elemental zinc was used orally alongside the zinc ionophore, hydroxychloroquine, but not when either intervention was used alone,^16^ and the addition of 100mg of elemental zinc to a hydroxychloroquine/azithromycin protocol was also found to reduce the risk of mortality or transfer to hospice care.^17^ Two other studies however, reported minimal, if any, benefit from zinc use on the survival of adults hospitalised with SAR-CoV-2 infection.^18 19^ Regarding community treatment, in a case series of four adults with confirmed or suspected SARS-CoV-2 infections, symptomatic recovery coincided with the administration of high dose zinc lozenges of 115mg to 207mg elemental zinc daily.^20^

Most covid-19 prevention and treatment guidelines are yet to mention zinc.^21-27^ An exception is the National Institute of Health (NIH) that in July 2020, stated there is insufficient evidence to make any recommendations for acute treatment with zinc.^28^ Based on expert opinion, the NIH made a moderately strong recommendation against the use of zinc in doses above the recommended daily intake of zinc for the prevention of covid-19 (i.e. 8mg to 14mg of elemental zinc for adults). This was due to concerns about secondary copper deficiency.^29 30^

The rationale for zinc has been reviewed in detail elsewhere.^5 31 32^ In summary, zinc has direct antiviral properties against SARS-CoV-2 and other coronaviruses.^31-33^ There is the potential for broader multisystem effects via modulation of angiotensin converting enzyme 2 activity, inflammation, immunity, haemostasis and hypoxic responses.^5 31 32 34^ Further, the risk factors for severe illness from SARS-CoV-2 infection, such as increasing age, obesity and chronic disease, are also risk factors for zinc deficiency.^5 32 35^ Observational studies conducted in Germany, Spain, Japan and Iran have reported that lower baseline zinc levels in hospitalised adults were associated with a higher risk of mortality, severe illness, complications, and longer hospital stay following SARS-CoV-2 infection.^33 36-38^

In early 2020, the World Naturopathic Federation responded to a World Health Organization initiative by calling for rapid evidence reviews to inform self-care and clinical practice during the covid-19 pandemic. We published a rapid review protocol to evaluate zinc for the prevention and treatment of SARS-CoV-2 and other viral respiratory tract infections (RTIs).^39 40^ The aim was to assess the effects of zinc on the incidence, duration and severity of acute SARS-CoV-2 infections in people of any age or zinc status, and to include the best available evidence. Indirect evidence was sought as the risk of other viral RTIs remains relevant during the pandemic, zinc is often self-prescribed or recommended prior to knowing the cause, and systematic reviews of zinc for non-specific RTIs were either outdated, limited by population or administration route, or low quality.^41-48^ Ongoing uncertainty makes it difficult to make informed decisions, either for or against the use of zinc for SARS-CoV-2 or other viral RTIs in populations who are not zinc deficient.^5 27^

The scope of randomised controlled trials (RCTs) involving people of any age, published in English or Chinese, along with the details of four RCT protocols evaluating the efficacy of zinc to prevent or treat SARS-CoV-2 infections in adults were reported in August 2020^5^. Of the 118 RCTs with indirect evidence, the 25 RCTs that included adult populations were prioritised for analysis, as this was an *a priori* subgroup population with a higher risk of severe acute respiratory syndrome (SARS).

This rapid review presents an updated search and meta-analysis of the RCTs investigating any type of zinc intervention to prevent or treat SARS-CoV-2 or other viral RTI in adult populations.

## METHODS

### Protocol

This rapid review (RR) conforms with the Interim Guidance from the Cochrane Rapid Reviews Methods Group,^2^ the Cochrane Handbook,^49^ and the Preferred Reporting Items for Systematic Reviews and Meta-Analyses (PRISMA)^50^. The protocol is registered with the International Prospective Register of Systematic Reviews, number CRD42020182044^39^. Following feedback from content experts, the protocol was updated.^40^ The inclusion criteria was expanded from only including respiratory infection likely to be caused by a coronavirus to those caused by any virus, the exclusion criteria was tightened so that included studies only included respiratory illnesses mostly caused by viral infections, and the planned database search was expanded. Post-protocol input from consumer/patient advocate representatives who were blinded to the results, led to minor changes to the outcomes and the rating of their importance (Table 1). The other post-protocol changes were the decisions to stagger the analysis and periodically update the review for direct SARS-CoV-2 evidence.

**Table 1.**
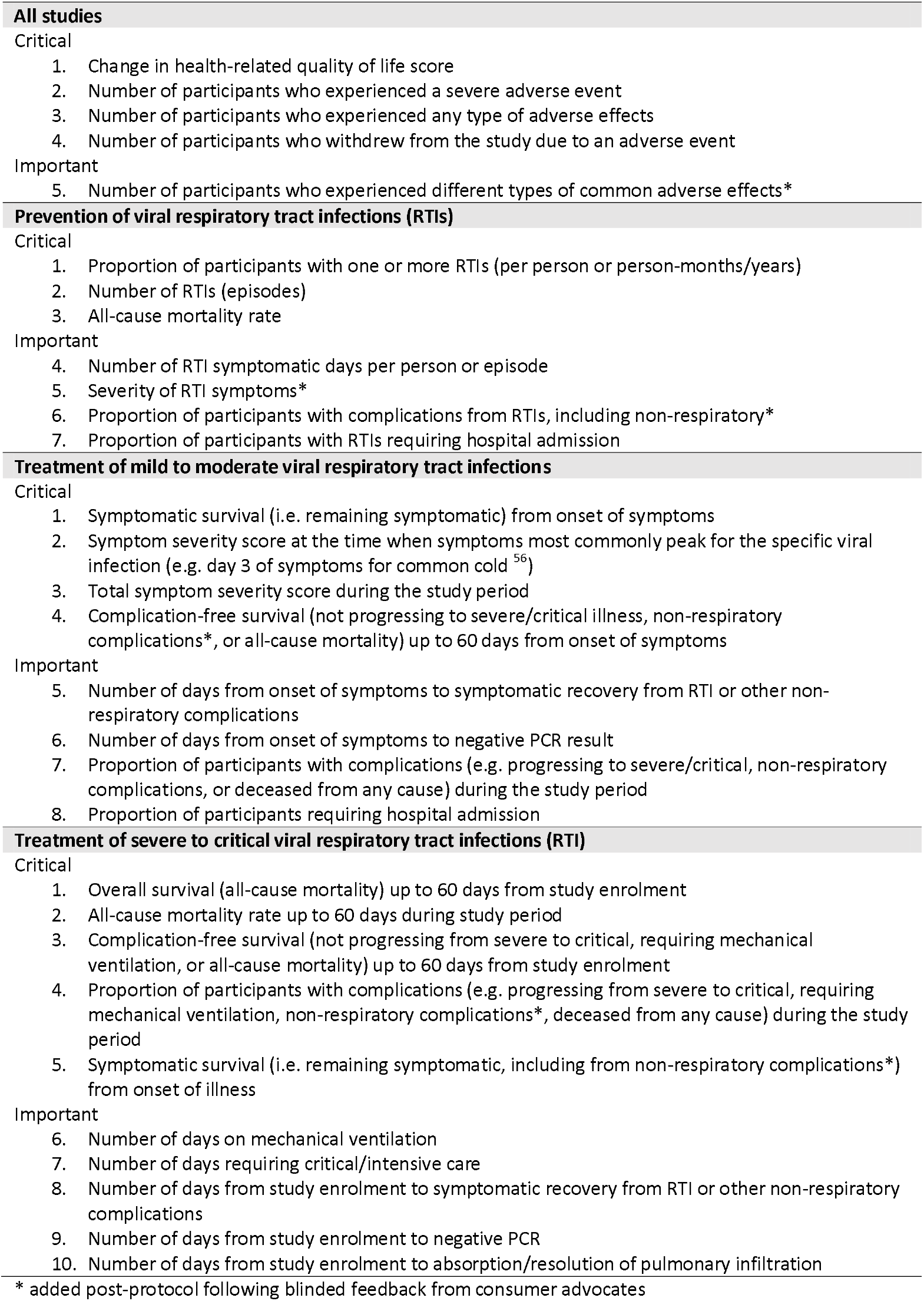
Critical and Important Outcomes.

### Search strategy

The search strategy was developed in collaboration with an experienced research librarian (JB). Subject headings and keywords were developed for coronaviruses, viral respiratory illnesses, zinc, and randomised controlled trials in humans. The following databases were searched with no limits on language nor date: PubMed, Embase, Cochrane CENTRAL, Academic Search Complete, Allied and Complementary Medicine Database, Alt Health Watch, CINAHL Plus with Full Text, Health Source, PsycINFO, China Knowledge Resource Integrated Database (CNKI), medRxiv, bioRxiv, U.S. National Library of Medicine Register of Clinical Trials (ClinicalTrials.gov), International Standard Randomized Controlled Trial Number Register (ISRCTN), World Health Organization International Clinical Trials Registry Platform (WHO ICTRP), Global Coronavirus covid-19 Clinical Trial Tracker and Chinese Clinical Trial Registry. Database searches were conducted between 29 April and 15 August 2020 and supplemented by bibliography searches of included articles (Appendix 1 – available upon request).

### Study selection criteria

#### Study design

Included were randomised and quasi-randomised controlled trials. Excluded were systematic reviews, non-randomised studies of interventions, and studies without a concurrent control.

#### Population

Direct evidence from adults in any setting who were at risk of, or had a laboratory confirmed SARS-CoV-2 infection. In anticipation of the likely dearth of direct evidence so early in the pandemic, indirect evidence from other viral respiratory tract infections (RTIs) were included due to zinc’s known mechanisms of action,^5^ and in response to calls for the best available evidence, even if indirect, to inform clinical decisions and research. As such, laboratory confirmed RTIs and non-specific respiratory tract illness predominantly caused by a viral infection, such as the common cold, non-seasonal rhino-sinusitis, pharyngitis, laryngitis, flu-like illness and healthy adults with acute bronchitis were included. Excluded were non-specific bronchitis in adults with concurrent chronic lung diseases, pneumonia, otitis and acute or chronic respiratory distress/failure. Studies with eligible and ineligible participants were included (e.g. adults or participants with viral RTIs) and when possible, only the data for the eligible population was extracted.

#### Interventions and comparators

Included were interventions of any zinc conjugates, dose, duration, and administration route. Excluded were co-interventions, including other nutraceuticals, herbs, or pharmaceuticals unless both the intervention and control groups received the co-intervention. All types of controls and comparator groups were included.

#### Outcomes

Critical and important outcomes were informed by core outcome sets,^51-54^ and a withdrawn Cochrane protocol for zinc for the common cold,^55^and prioritised based on their importance to patients and healthcare practitioners (Table 1). Studies were included regardless of the outcomes reported and whether these were primary or secondary outcomes. Outcomes not of interest were noted but not analysed.

### Data collection and appraisal

In line with recommended RR methods,^2^ the first 30 title-abstracts and 5 full-papers were jointly screened for calibration and consistency, the remaining were screened by single, experienced reviewers. A high threshold for exclusion was applied and all studies excluded at full-paper screen were rescreened by a second reviewer. Following calibration, a single reviewer extracted the data and appraised the risk of bias that was verified by second reviewer. The exceptions were articles published in Chinese where screening, data extraction and risk of bias appraisal were conducted by a single reviewer and discussed with the other reviewers. Other review constraints included not contacting study authors for further information.

Study design and funding, participants, interventions, comparators, outcomes measures, effect size and direction were extracted into a piloted electronic spreadsheet. Data from graphical reports were extracted with WebPlotDigitizer Version 4.2^57^ with any rounding of decimal places and whole person estimates favouring the null hypothesis.

Quality was appraised with the revised Cochrane risk of bias 2.0 tool (Rob-2).^58^ RCTs published prior to 2002 were not penalised for not publishing a protocol. Rapid review constraints included only appraising the outcomes that were meta-analysed. If no outcomes were analysed, the study’s primary outcome was appraised. Only one reviewer appraised an outcome according to a pre-piloted rubric. A seconder reviewer verified the appraisal with disagreements resolved through consensus.

### Evidence synthesis and statistical methods

The effect measures for dichotomous outcomes were risk ratios, calculated using the Mantel-Haenszel method. Events measured over different timeframes were calculated and reported as the incidence rate per person-months, from which rate ratios and rate differences were estimated using an inverse variance method. Studies reporting separate counts for different types of viral RTIs (e.g. common cold, bronchitis, flu-like illness) were combined to calculate the total number of RTIs. When no RTIs were reported in one study arm, 0.5 was recorded to facilitate analysis. For continuous data, either the weighted or standardised mean differences were calculated using an inverse variance method. Mean symptom severity scores were transformed to a modified Jackson common cold scale.^59^ For time-to-event data, hazard ratios (HR) were calculated using an <u>O</u>-E and variance method. Data extracted from survival curves was imputed for the first seven days using the direct method 10 in the ‘HR calculations spreadsheet’ published by Tierney et al.^60^ For the purpose of estimating an absolute effect from zinc use, the probability of remaining symptomatic on day-7 was set at 33% for the placebo and active control comparators.^56^

Review constraints for non-SARS-CoV-2 RCTs included not imputing missing data for any outcome, not imputing means or standard deviation (SD) for non-critical outcomes, and not contacting the authors. Instead, additional information from previous systematic reviews was used.^41-43^

Due to the large number of RCTs that assessed symptom severity yet could not be included in the meta-analyses, a basic count of the number of studies reporting significant and non-significant findings were narrated to provide further context.

RevMan 5.4,^61^ R software,^62^ Microsoft Excel, and GRADEpro GDT^63^ were used for the statistical analyses. Clinical and methodological diversity and statistical heterogeneity were considered prior to pooling two or more studies reporting a measure of effect.^64^ The random-effects model was used due to clinical and methodological diversity across the included studies. Statistical heterogeneity was assessed using the I^2^ statistic and homogeneity assessed with the chi^2^ test.

A *priori* subgroup analyses were conducted for different ages groups, causes and severity of RTIs and zinc interventions. Zinc doses were converted to milligrams (mg) of elemental zinc per day. To investigate potential dose effects of oral or sublingual zinc, the chi^2^ test comparing three categories (<50mg daily, 50-200mg daily, >200mg daily) was used for dichotomous and time-to-event data. The categories were based on a no observed adverse effect level (NOAEL) of 50mg and a higher risk of more severe adverse effects, such as vomiting, with doses above 225mg.^30^

The Grading of Recommendations Assessment, Development and Evaluation (GRADE) approach was used to rate the certainty of the estimated effects and reported in Summary of Findings (SoF) tables.^65^ Certainty was downgraded for indirectness by one level for SARS-CoV-1, MERS-CoV and other coronaviruses, and two levels for non-specific RTIs and other viral infections. Sensitivity analyses investigated the point estimate change of significant results when studies with a higher RoB or statistical outliers were removed, according to maximum days symptomatic prior to study enrolment or different definitions of symptomatic recovery, and when an alternate method 11 was used instead of method 10^60^ to impute the hazard ratios for individual studies. These sub-group and sensitivity analyses were used to assess the degree to which statistical heterogeneity might be explained by clinical or methodological diversity. When rating imprecision, the optimal information size of effect estimates was based on single-study sample size calculations of included studies, or a conventional 2-sided or equivalence sample size calculation, with an 80% power and a type 1 error rate of 5%. When data from at least 10 studies were pooled, funnel plots were created, visually inspected for publication bias and statistically analysed using Egger’s regression for continuous outcomes and the Harbord score for dichotomous outcomes.^66^

The minimally important difference (MID) in symptom severity for mild RTIs on day-3 was set at 1 point on a standardized scale (Appendix 4 – available upon request). This was the half-way mark between two proposed MIDs. Turner et al.^67^ proposed a 10% improvement for mild RTIs. Based on the pooled mean scores for the control arms on day-3, a MID would be 0.5 points. Norman et al.^68^ proposed that for populations with at least moderately impaired quality of life scores, the MID is approximately half the pooled standard deviation (SD) from the control arms, which for day-3 symptom severity would be 1.5 points. For standardised mean differences (SMD) the MID was set at 0.5 and a large effect size was 0.8.^69^ For duration of illness, a MID was at least twice as many participants recovering^70^ or one day shorter duration.^67^

### Patient and public involvement

The protocol was rapidly developed and published prior to patient advocate involvement. Experienced patient advocates based in Australia have since provided input, including blinded feedback on the importance of the outcome measures, and provided feedback on the presentation of the results and discussion. Ongoing involvement will include translating the review findings for consumers.

## RESULTS

### Included studies

From the 1,768 articles and registered trials retrieved through the database searches, 28 unique RCTs reported in 25 articles met the inclusion criteria (Figure 1).^67 70-93^ Three were published in Chinese language only.^90-92^ Appendix 2 is available upon request and lists the 95 RCTs evaluating zinc in paediatric populations, articles published in English that were excluded at full-paper screen, and the characteristics of the seven registered RCTs evaluating zinc for SARS-CoV-2 with pending results (registration numbers NCT04342728, ACTRN12620000454976, NCT04377646, PACTR202005622389003, IRCT20180425039414N2, NCT04447534 and CTRI/2020/07/026340).

**FIGURE 1.**
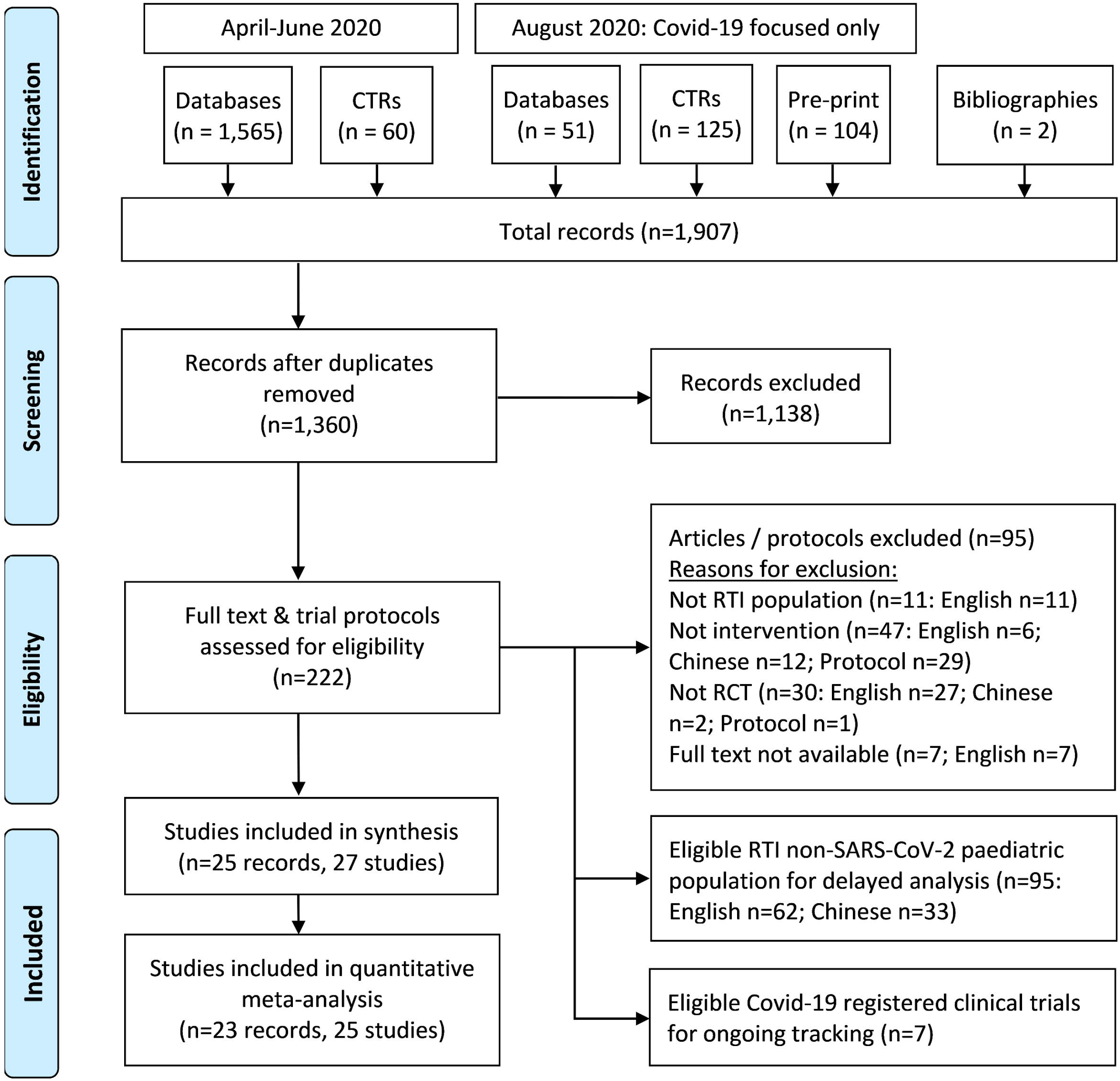
Search Results Flow Chart.

The characteristics of included studies are summarised in Table 2. Most were single-centre, 2-arm RCTs and were conducted in the United States (US). None of the RCTs included participants at risk of, or with a SARS-CoV-2 infection. Twenty RCTs evaluated zinc for community acquired infections that were mild to moderate severity ^67 70 75-93^ Six RCTs inoculated participants with a human rhinovirus strain (HRV 2, 13, 23 or 39).^67 72-74^ Two RCTs only assessed tolerability and adverse effects of a zinc lozenge designed to prevent or treat RTI.^71 72^

**Table 2.**
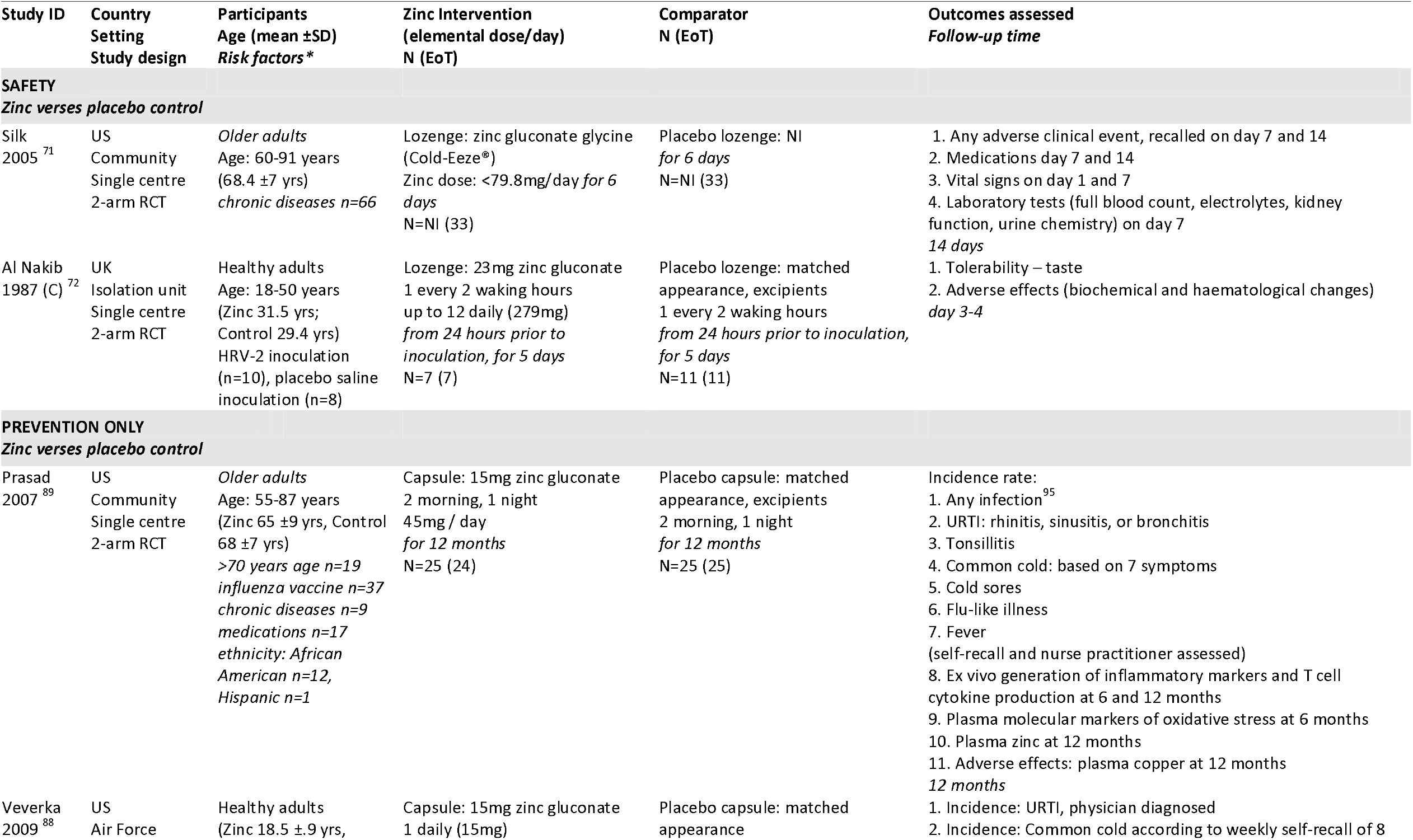
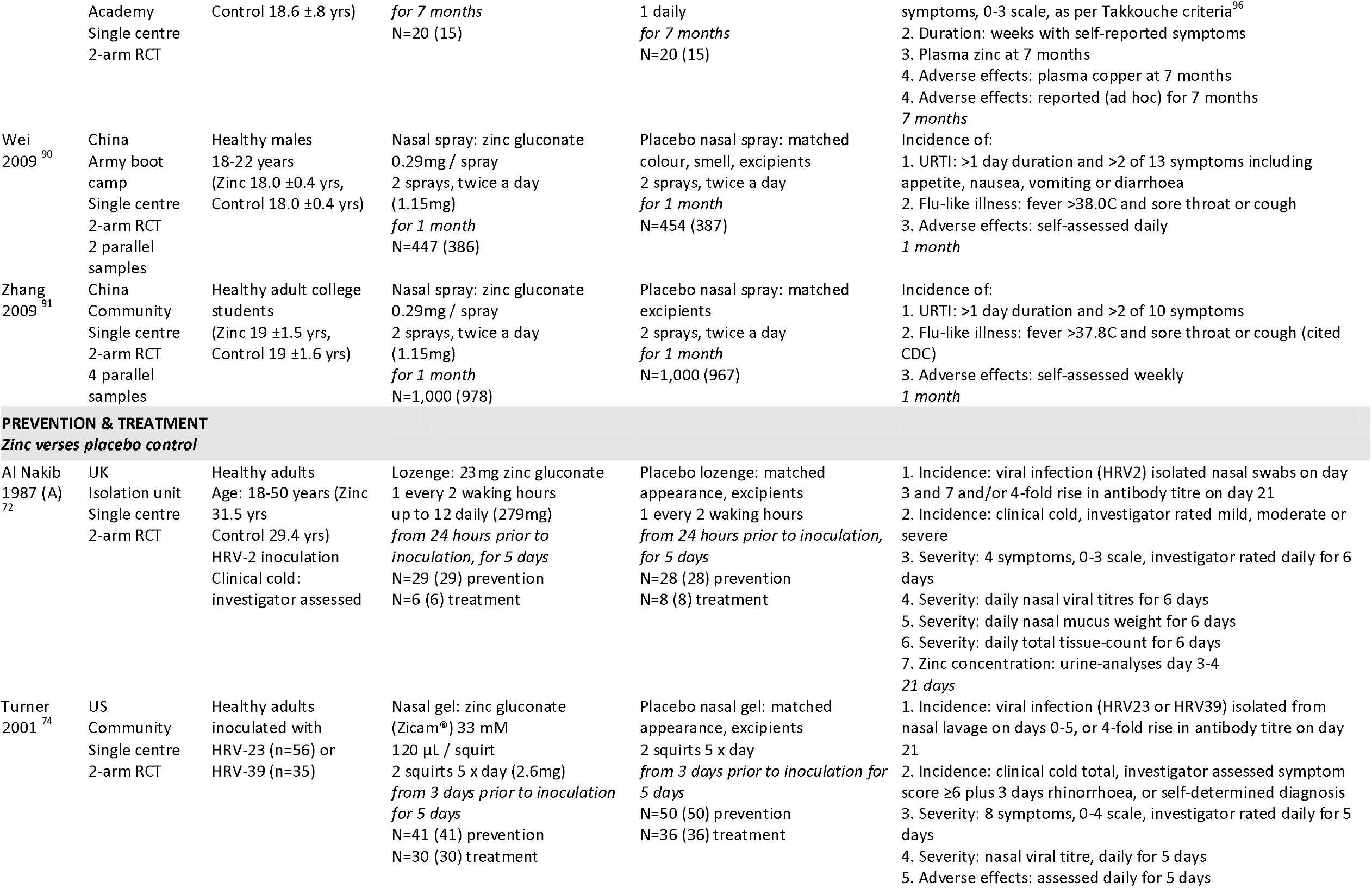
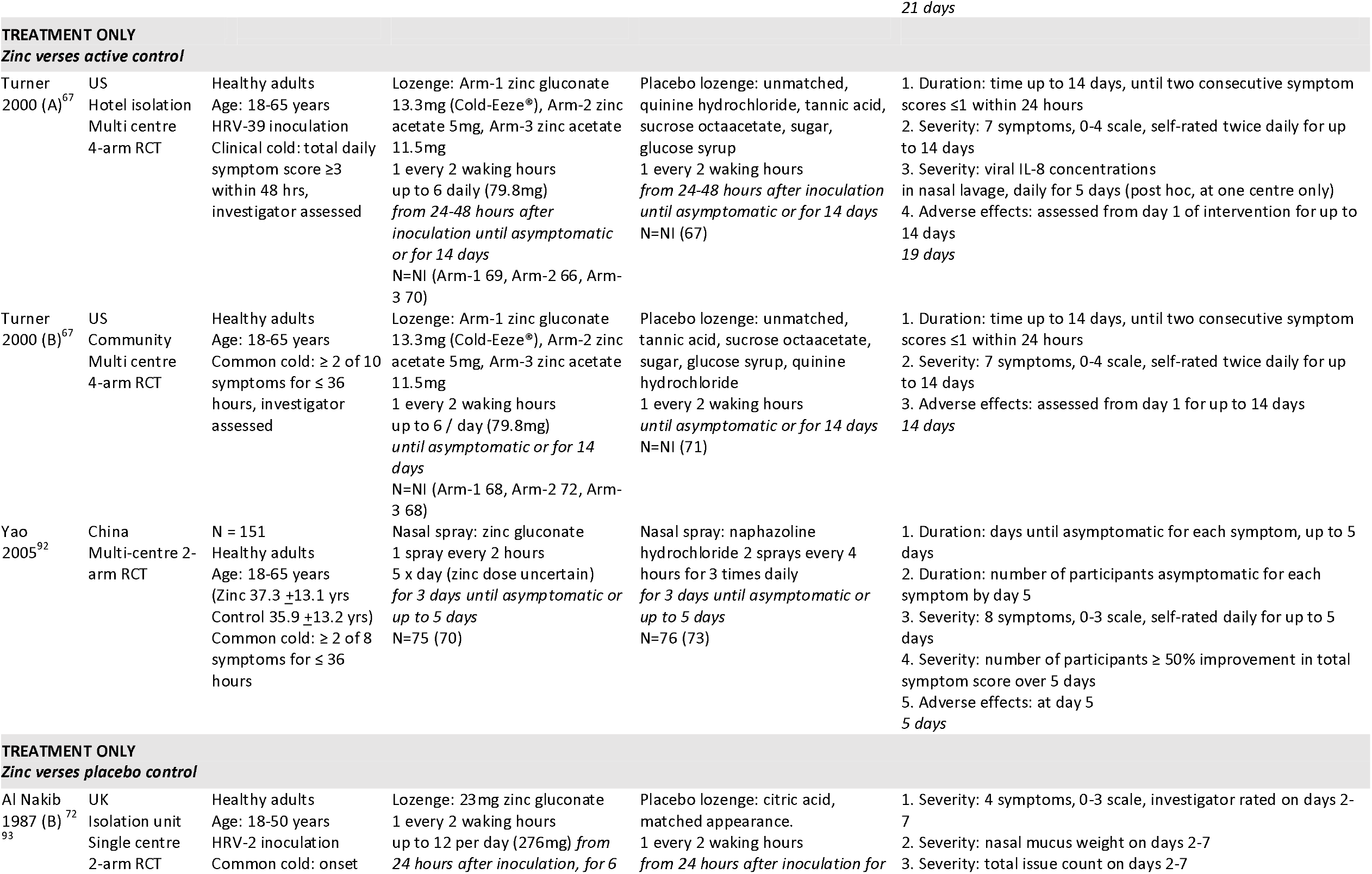
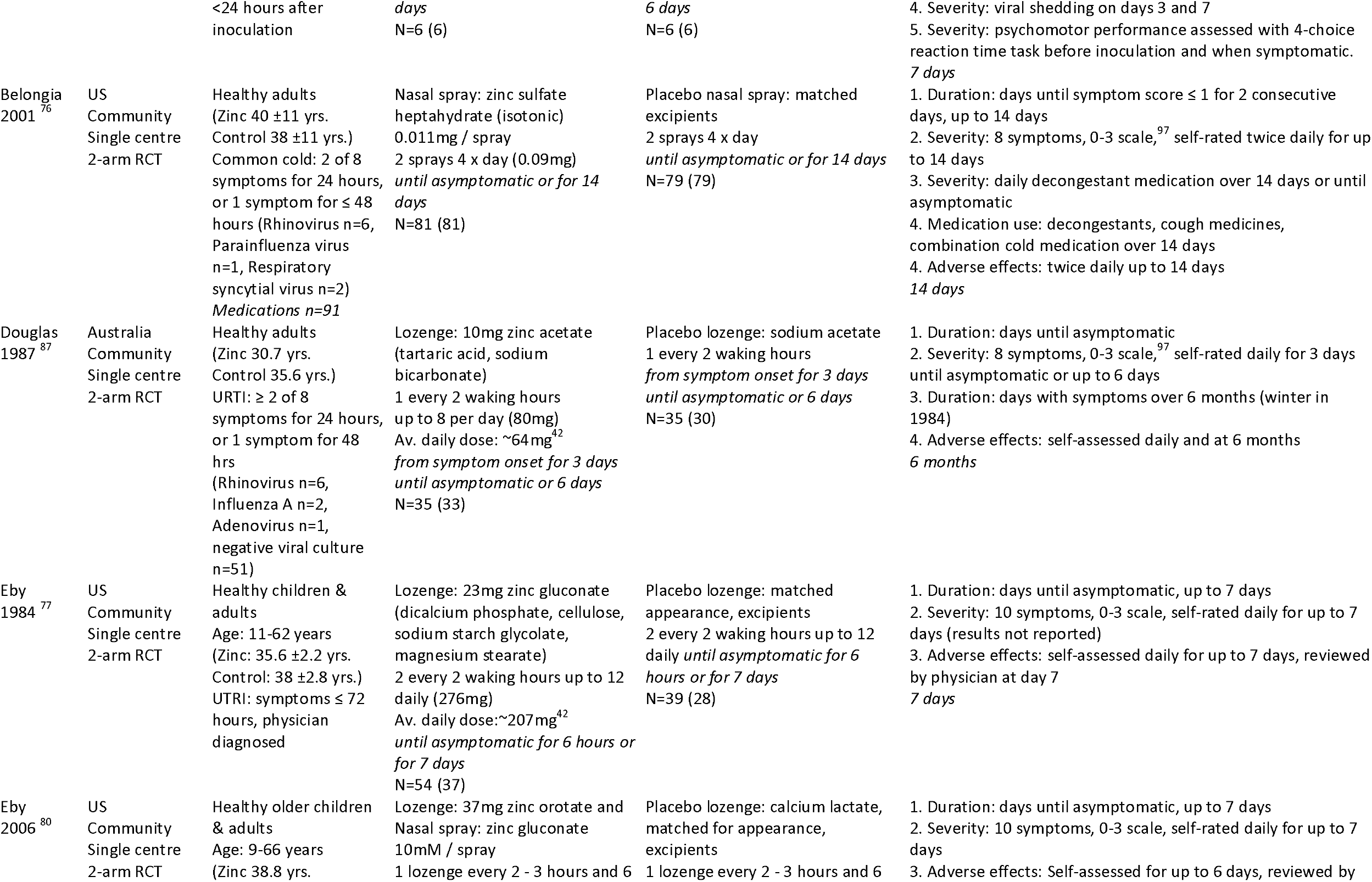
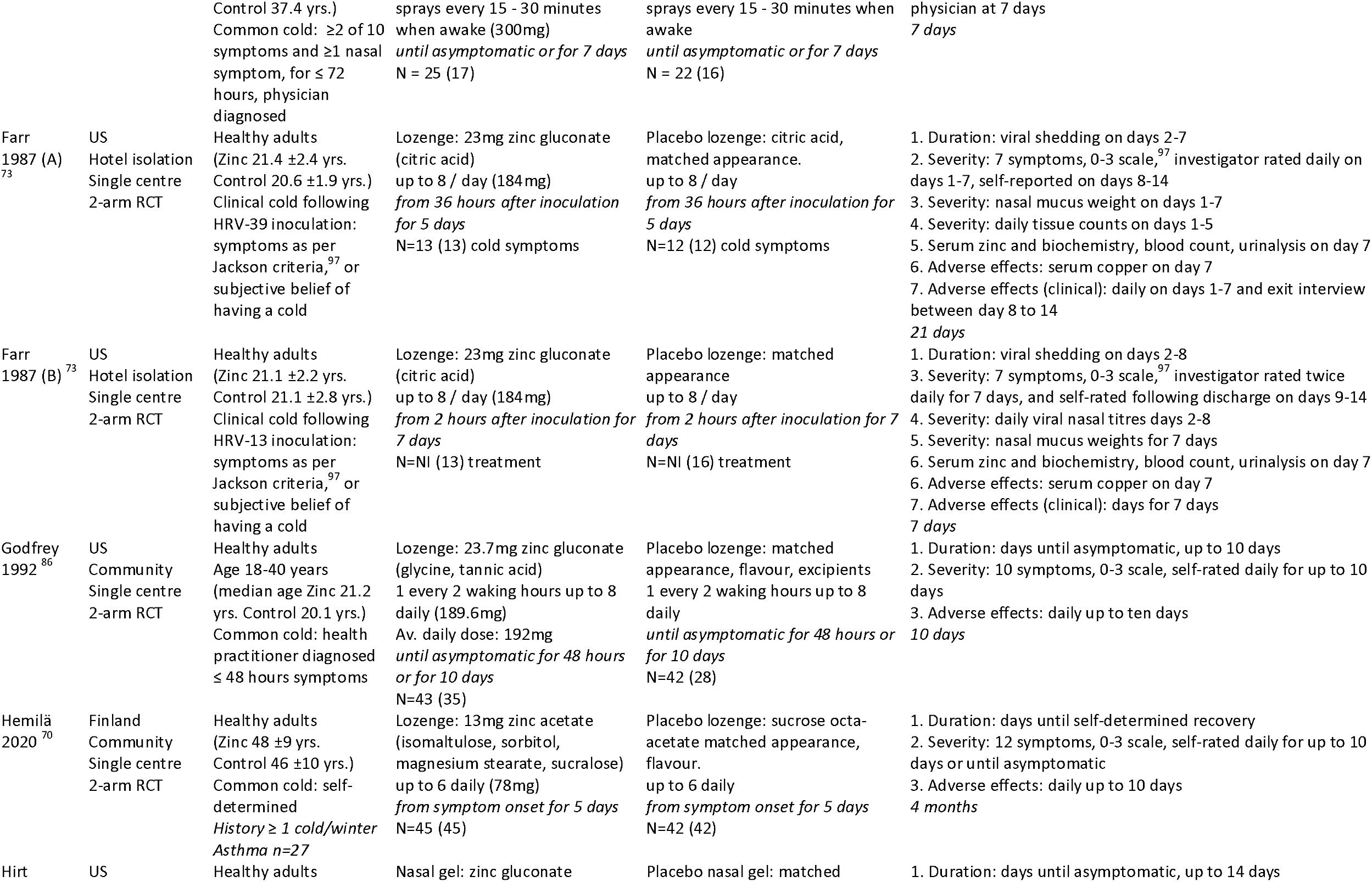
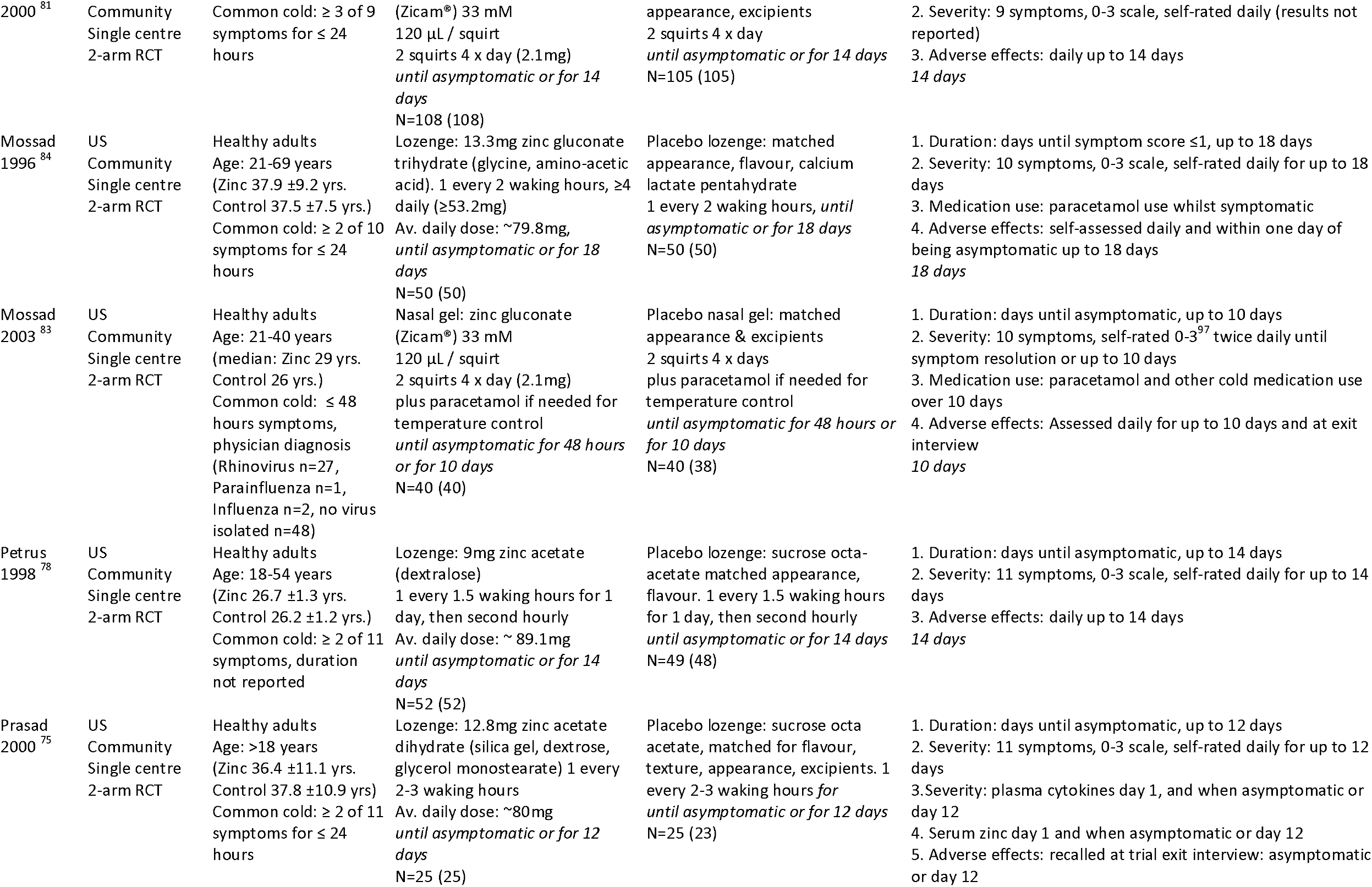
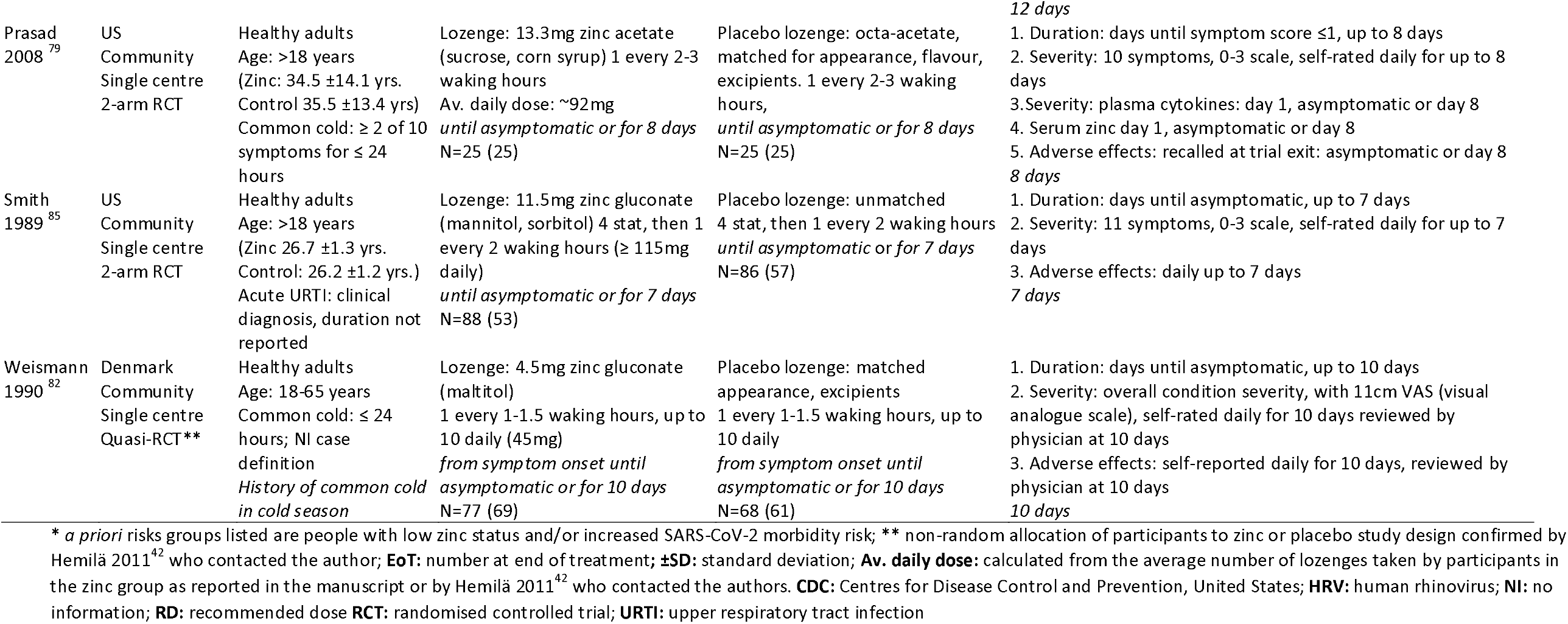
Characteristics of studies.

Most participants were younger than 65 years, with no SARS-CoV-2 risk factors (Table 2). However, two RCTs included older adults from different ethnic backgrounds, many of whom had chronic disease comorbidities and were taking long-term medication.^71 89^ In another RCT, around one third of participants had a history of asthma.^70^

The most common zinc formulations were lozenges followed by nasal sprays and gels containing either zinc acetate or gluconate salts (Table 2). The daily dose of elemental zinc from sublingual lozenges and oral capsules ranged from 15mg up to 300mg, whereas the doses for topical nasal applications were substantially lower (0.9-2.6 mg/day). Twenty-five RCTs compared zinc against a placebo that was often matched or partially matched. Two 4-arm RCTs described the control lozenge as a placebo,^74^ however, they were re-classified as an active comparator because they contained quinine hydrochloride that has broad-spectrum antiviral effects. ^94^Two RCTs permitted the concurrent use of paracetamol for temperature control and other cold medications for symptomatic relief.^76 83^ The use of breakthrough medication was reported and applied to all study participants. All but two RCTs ^71 72^ reported at least one result that was used in a meta-analysis of a critical or important outcome. None of the RCTs reported mortality nor quality of life outcomes. The details of the meta-analyses, subgroup and sensitivity analyses, funnel plots, and any additional calculations used for the analysis can be found in Appendix 4 – available upon request.

The certainty of the evidence for all outcome measures was downgraded for SARS-CoV-2 indirectness (Table 3). A high risk of bias for some outcomes, inconsistency, and imprecision additionally undermined confidence. Concerns about the risk of bias included probable unmasking due to the challenges with matching the placebo and higher rates of adverse effects from zinc formulations such as nausea, taste changes and mouth or nasal irritation that could influence participant reported outcomes (Figure 2 and Appendix 3 – available upon request). For symptom severity, insufficient reporting on missing responses to the daily questionnaire amplified other concerns about missing outcome data. Substantial heterogeneity lead to both measures of effect for duration of illness being downgraded. At least 11 RCTs were industry funded, with a further seven receiving partial industry support (Appendix 3 – available upon request). There was no obvious evidence of publication bias (Appendix 4 – available upon request).

**Table 3.**
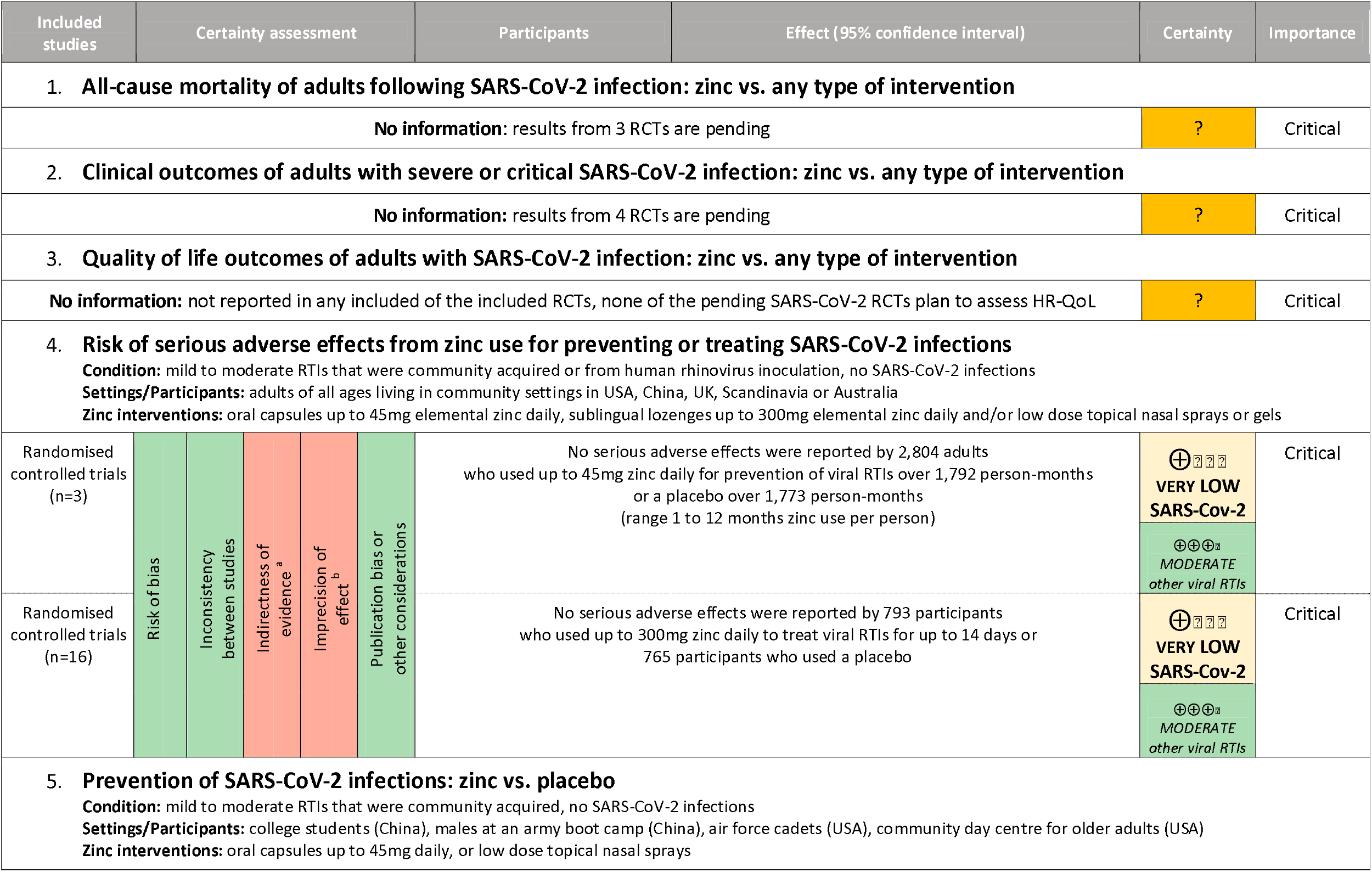
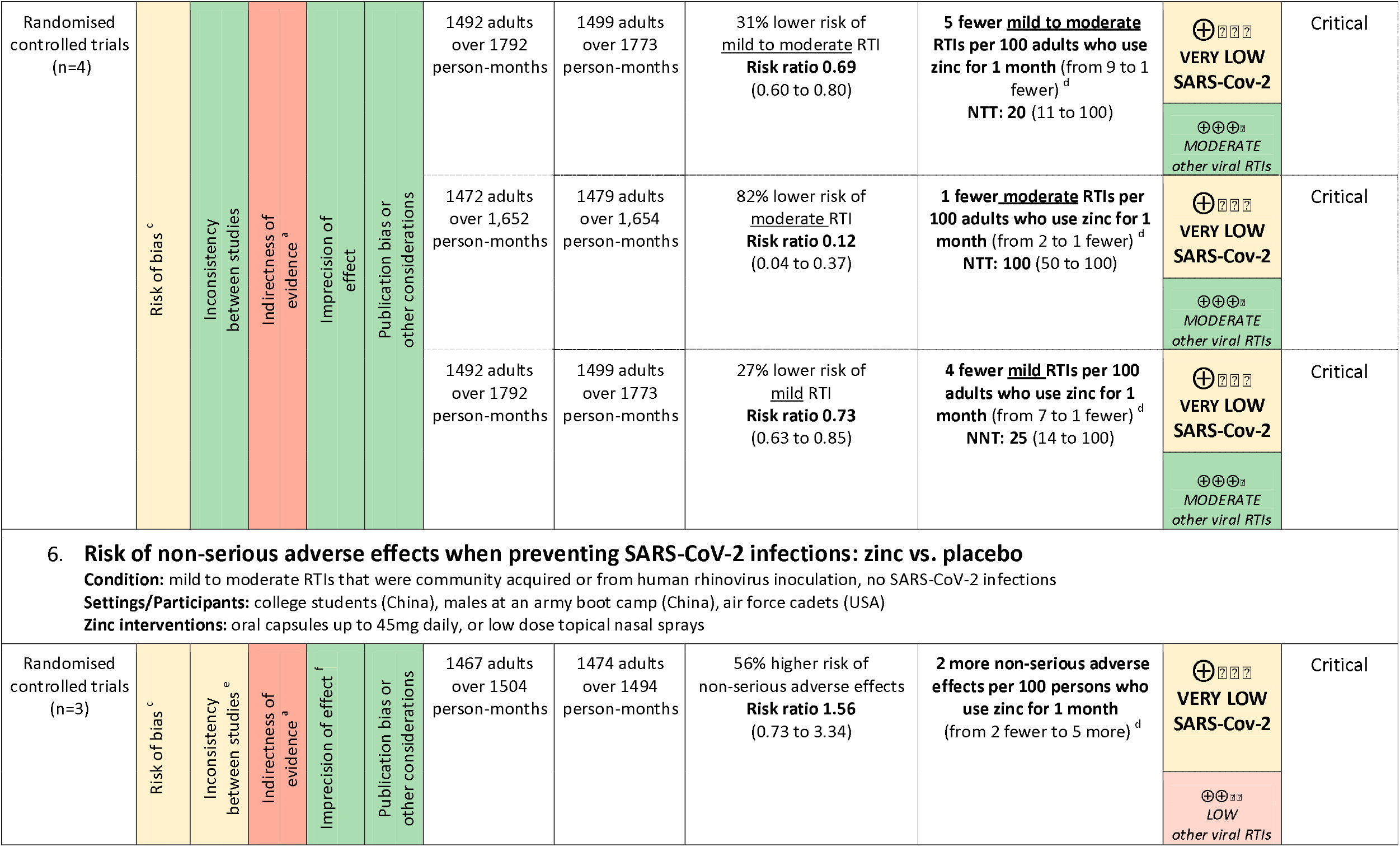
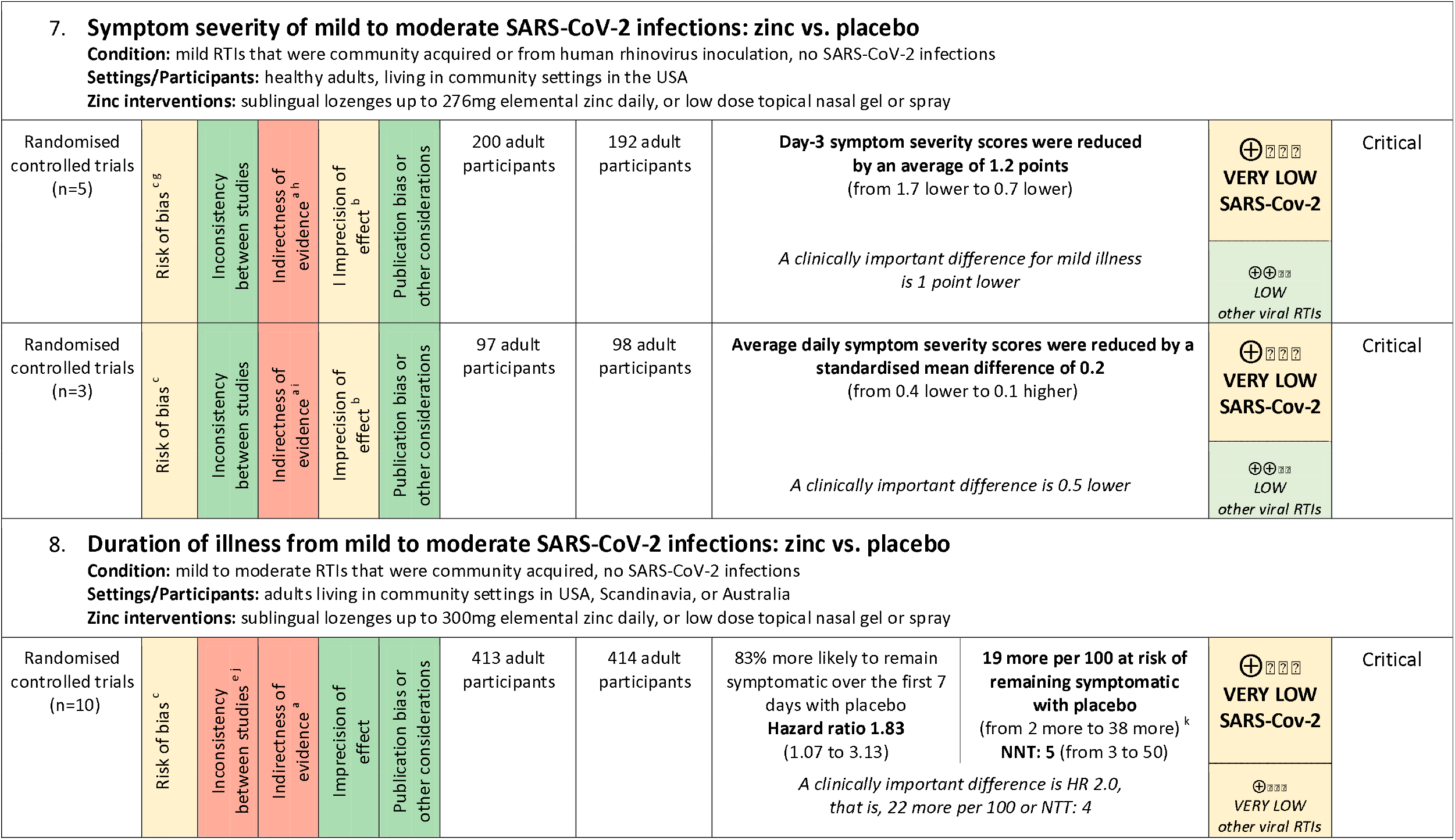
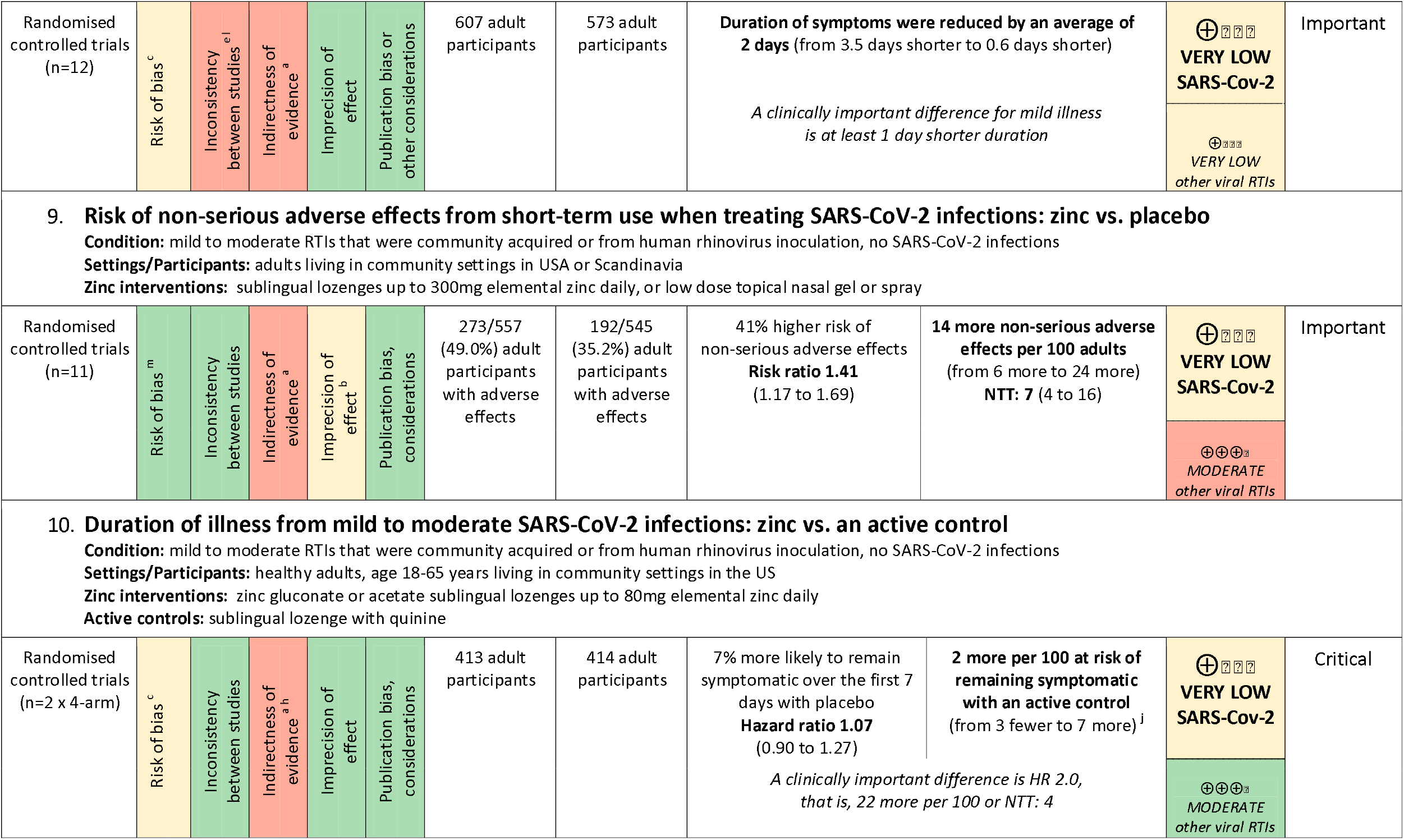
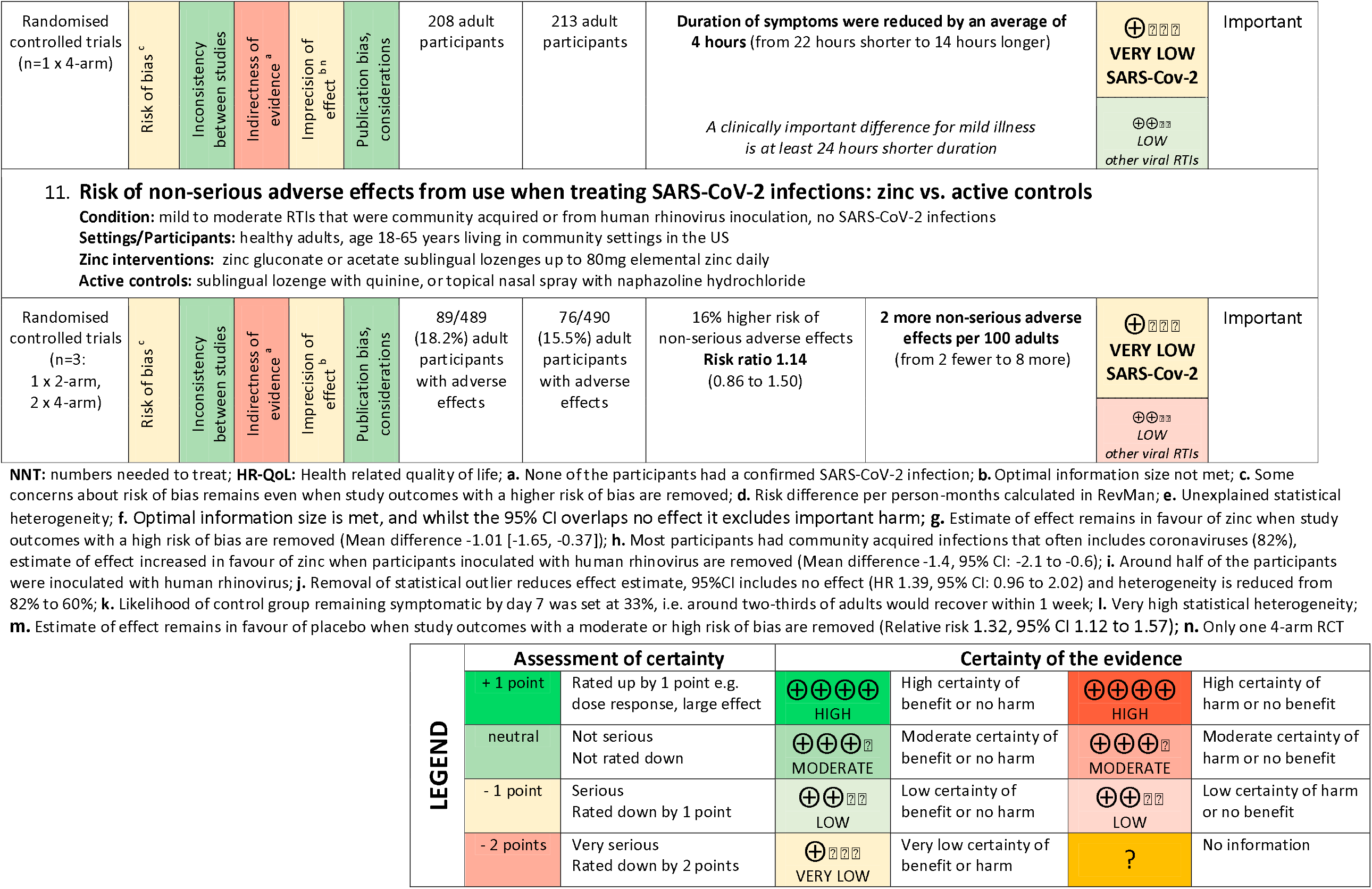
Summary of Findings: efficacy and safety of zinc interventions for preventing or treating SARS-CoV-2 infections in adults.

**FIGURE 2.**
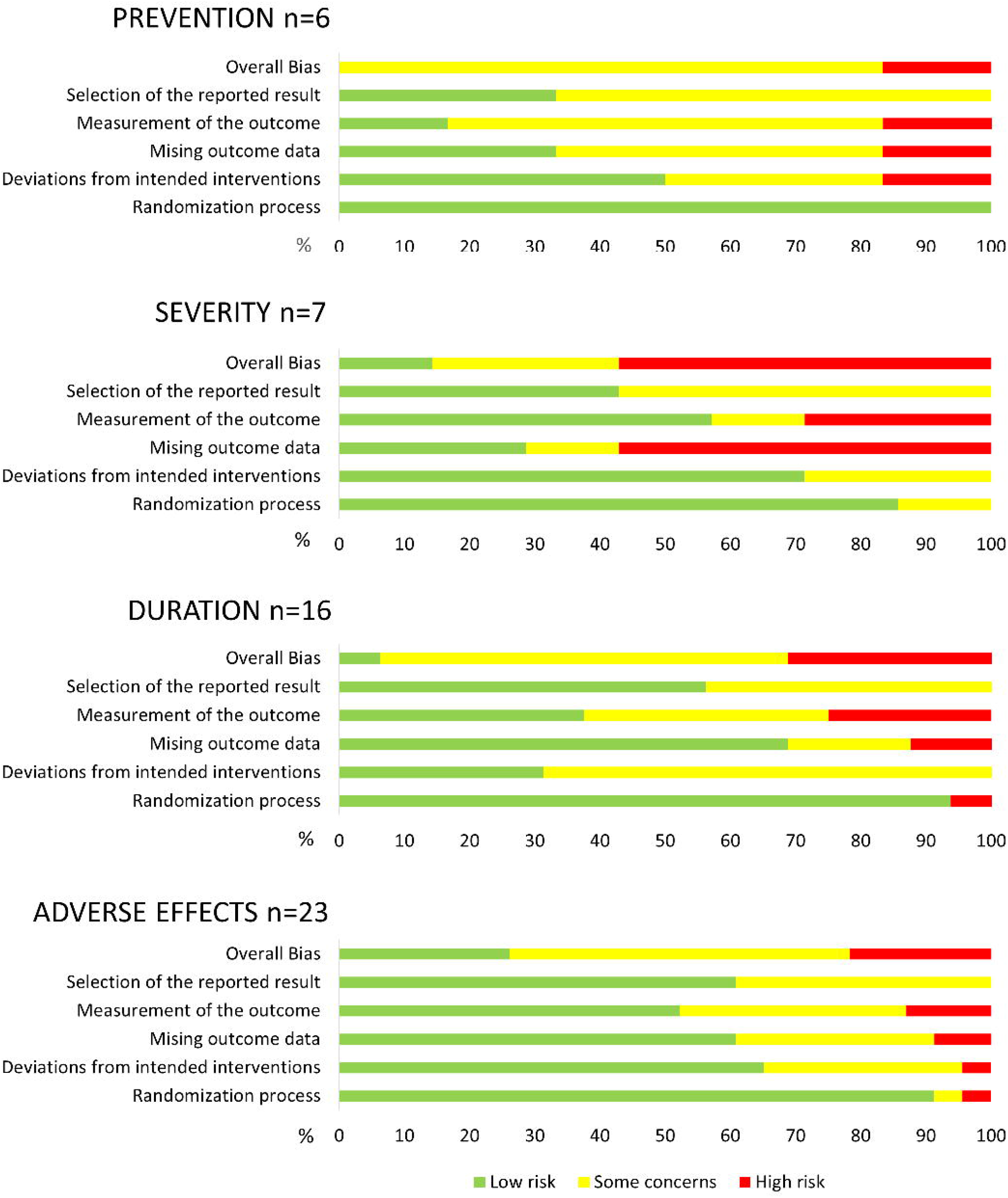
Risk of Bias.

### Zinc for prevention of SARS-CoV-2 infection

There were no data specific to SARS-CoV-2 nor other coronaviruses, consequently, confidence in all estimated effects was downgraded to very low certainty (Table 3).

#### Community acquired infections

Prevention of community acquired viral RTIs was investigated in four RCTs that included 2,804 participants over a total of 3,565 person-months.^88-91^ Compared with placebo controls, zinc reduced the risk of developing symptoms consistent with a mild to moderate RTI by 31% (Risk Ratio (RR) 0.69, 95% Confidence Interval (CI) 0.60 to 0.80, very low certainty SARS-CoV-2, moderate certainty for other RTIs) (Figure 3). The number needed to treat (NTT) to prevent an RTI was 20 adults using zinc for one month (95% CI 11 to 100). The largest effect was an 88% reduction for moderately severe, flu-like illnesses (RR 0.12, 95% CI 0.04 to 0.37, very low certainty SARS-CoV-2, moderate certainty other RTIs).^88-91^ However, due to the higher incidence of mild RTIs (e.g. common cold), even with a lower risk reduction of 29% (RR 0.71, 95% CI 0.60 to 0.80) the absolute effect was larger (Table 3).^88-91^Four mild RTIs (95% CI 7 to 1) might be prevented with 100 person-months of zinc use, compared to one moderately severe RTI (95% CI 2 to 1). For the subgroup of older adults aged 55-87 years, based on one RCT only (49 participants, 588 per-person months), the risk of RTIs was more than halved (RR 0.38, 95% CI 0.16 to 0.90, very low certainty SARS-CoV-2, low certainty other RTIs).^89^ Subgroup analysis found no significant differences according to zinc administration route or dose (P=0.37) (Figure 3).

**FIGURE 3.**
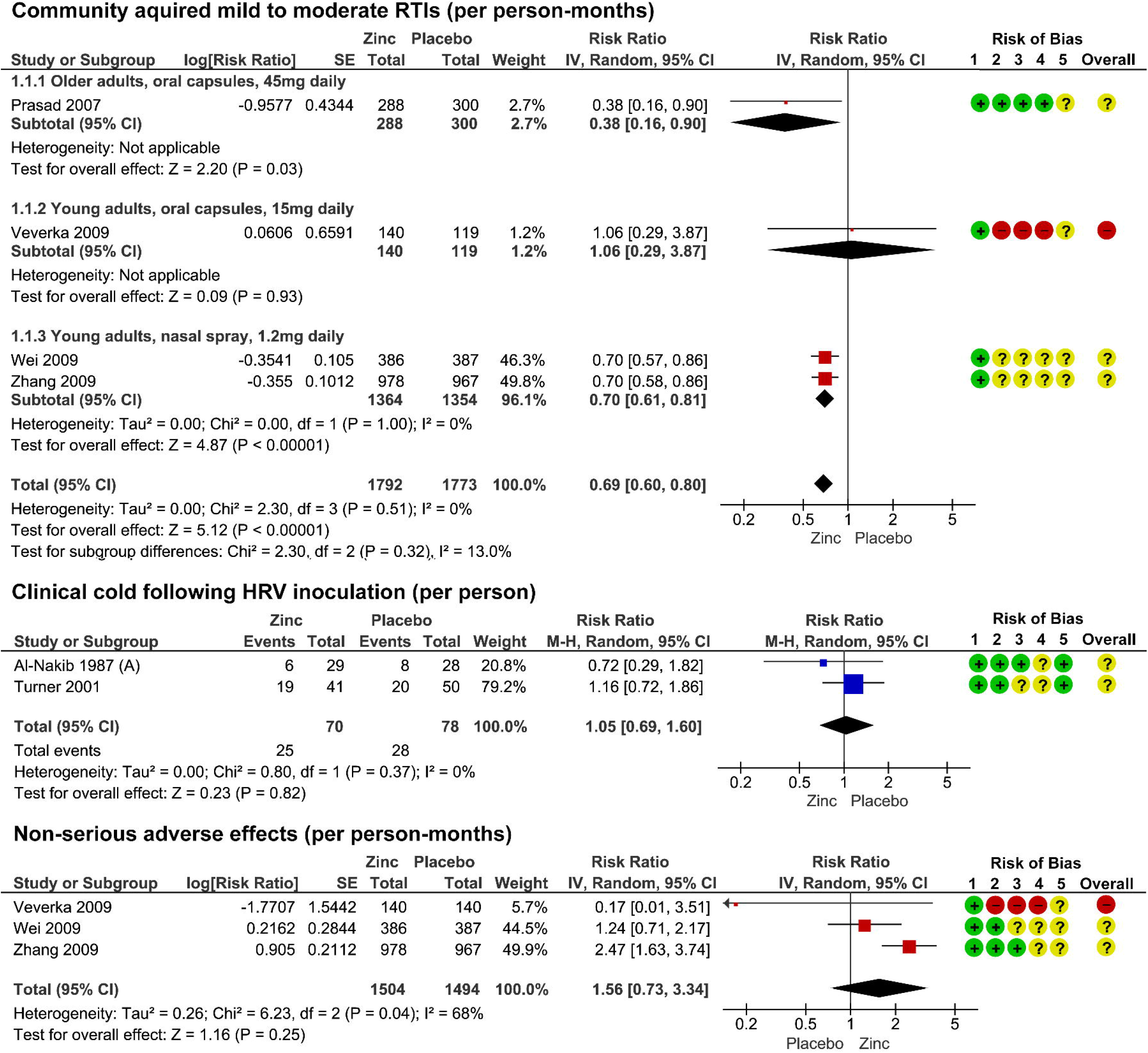
Prevention of RTIs and adverse effects.

#### Human rhinovirus inoculation

The effect of zinc compared to placebo for preventing RTIs symptoms following inoculation with human rhinoviruses was evaluated in two RCTs that included 148 participants.^72 74^ Neither study found a difference in the risk of infection nor in developing a clinical cold (RR 1.05, 95% CI 0.69 to 1.60, very low certainty SARS-CoV-2 and other RTIs) (Figure 3).

#### Adverse effects

No serious adverse events were reported (very low certainty SARS-CoV-2, moderate certainty other RTIs). Anosmia (loss of sense of smell) was not reported by the 1,447 participants who used a zinc nasal spray nor the 1,354 participants who used a placebo spray for 1 month (very low certainty SARS-CoV-2, moderate certainty other RTIs).^90 91^ Compared to placebo, no differences in copper plasma concentration were found in the two smaller RCTs that evaluated 15mg of oral zinc for younger adults over 7 months^88^ or 45mg for older adults over twelve months^89^ (very low certainty SARS-CoV-2, low certainty other RTIs). Based on the pooled results of three RCTs that included 2,758 participants and 2,998 per person-months, there were no differences in the risk non-serious adverse effects from zinc compared to placebo controls (RR: 1.56, 95% CI 0.73 to 3.34, very low certainty SARS-CoV-2, low certainty other RTIs) (Figure 3).^88 90 91^

### Zinc for treatment of SARS-CoV-2

There were no data specific to SARS-CoV-2 nor other specific coronaviruses, consequently, confidence in all estimated effects was downgraded to very low certainty for SARS-CoV-2 (Table 3).

#### Symptom severity

Of the 23 RCTs that evaluated the effects of zinc on symptom severity for non-specific and other viral RTIs,^67 70 72^-^87 92^ only seven reported results that could be extracted for a meta-analysis of an *a priori* outcome (Figure 4).^73-79^

**FIGURE 4.**
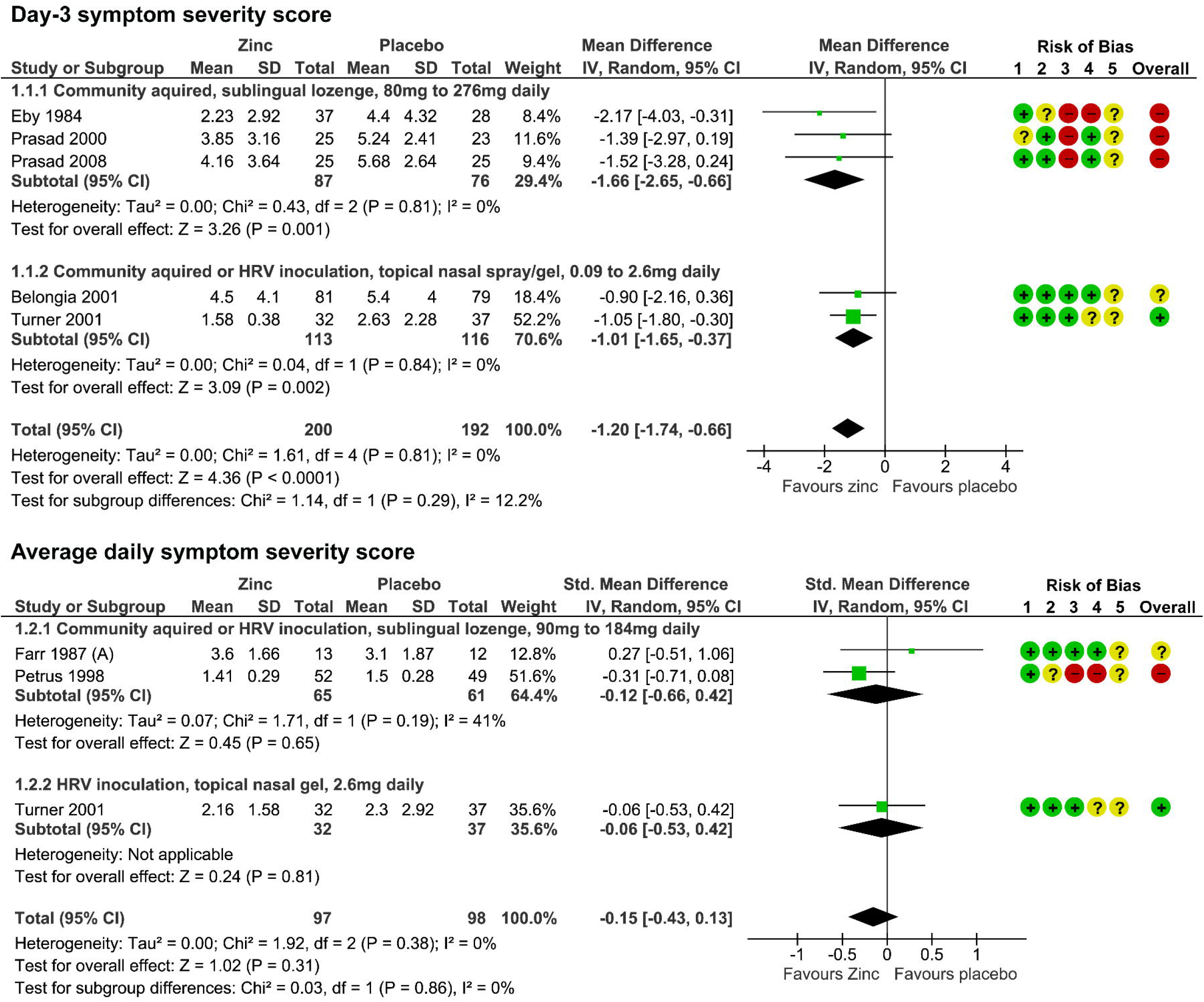
Symptom severity.

In five RCTs that included 392 participants, a clinically important reduction of more than 1 point in the day-3 symptom severity scores for mild RTIs was found for zinc compared to placebo (mean difference (MD) −1.21, 95% CI −1.74 to −0.66, very low certainty SARS-CoV-2, low certainty for other RTIs).^74-77 79^ Subgroup analyses found no significant differences between zinc administered as a topical nasal spray or gel compared to sublingual lozenges, nor were there any differences between RTIs that were community acquired compared to those caused by rhinovirus inoculation.

Average daily symptom severity scores were no different for zinc compared to placebo controls (standardized mean difference (SMD −0.15, 95% CI −0.43 to 0.13, very low certainty SARS-CoV-2, low certainty other RTIs) in a pooled analyses of three RCTs, including a total of 195 participants.^73 74 78^ Similarly, the effect for zinc compared to placebo was unchanged by its administration route and by the type of viral infection.

In two multicentre 4-arm RCTs, three different zinc lozenges were compared to an active control containing quinine hydrochloride. In both RCTs, one with 272 participants who were inoculated with human rhinovirus and the other with 279 participants who had a community acquired, non-specific mild viral RTI, symptom severity was equivalent.^67^ Similarly, in another RCT that included 143 participants,^92^symptom severity with zinc nasal spray use was equivalent to an active control nasal spray containing naphazoline hydrochloride.

Of the remaining RCTs that evaluated the effects of zinc compared to placebo on symptom severity according to various outcome measures, five RCTs including a total of 353 participants, reported lower symptom severity from zinc,^72 83-86^ four RCTs that included 248 participants, reported no differences between groups,^72 73 82 87^ and three RCTs did not report their results.^70 80 81^

#### Duration of illness

Zinc was found to effectively reduce the duration of community acquired mild to moderately severe RTIs in 14 RCTs that included 1,300 participants (Figure 5). ^70 75-87^ The pooled results from 10 RCTs that followed up 1023 participants for at least seven days,^70 75-77 80-85^ found that on any given day, the risk of remaining symptomatic was 83% higher for placebo controls compared to zinc (HR 1.83, 95% CI 1.07 to 3.13, very low certainty SARS-CoV-2 and other RTIs). The pooled results from 12 RCTs that included 1,180 participants,^75-79 81-87^ found the duration of symptoms was reduced by an average of two days with zinc compared to placebo controls (MD −2.03, 95% CI −3.50 to −0.59, very low certainty SARS-CoV-2 and other RTIs), more than twice the minimally important difference of one day.

**FIGURE 5.**
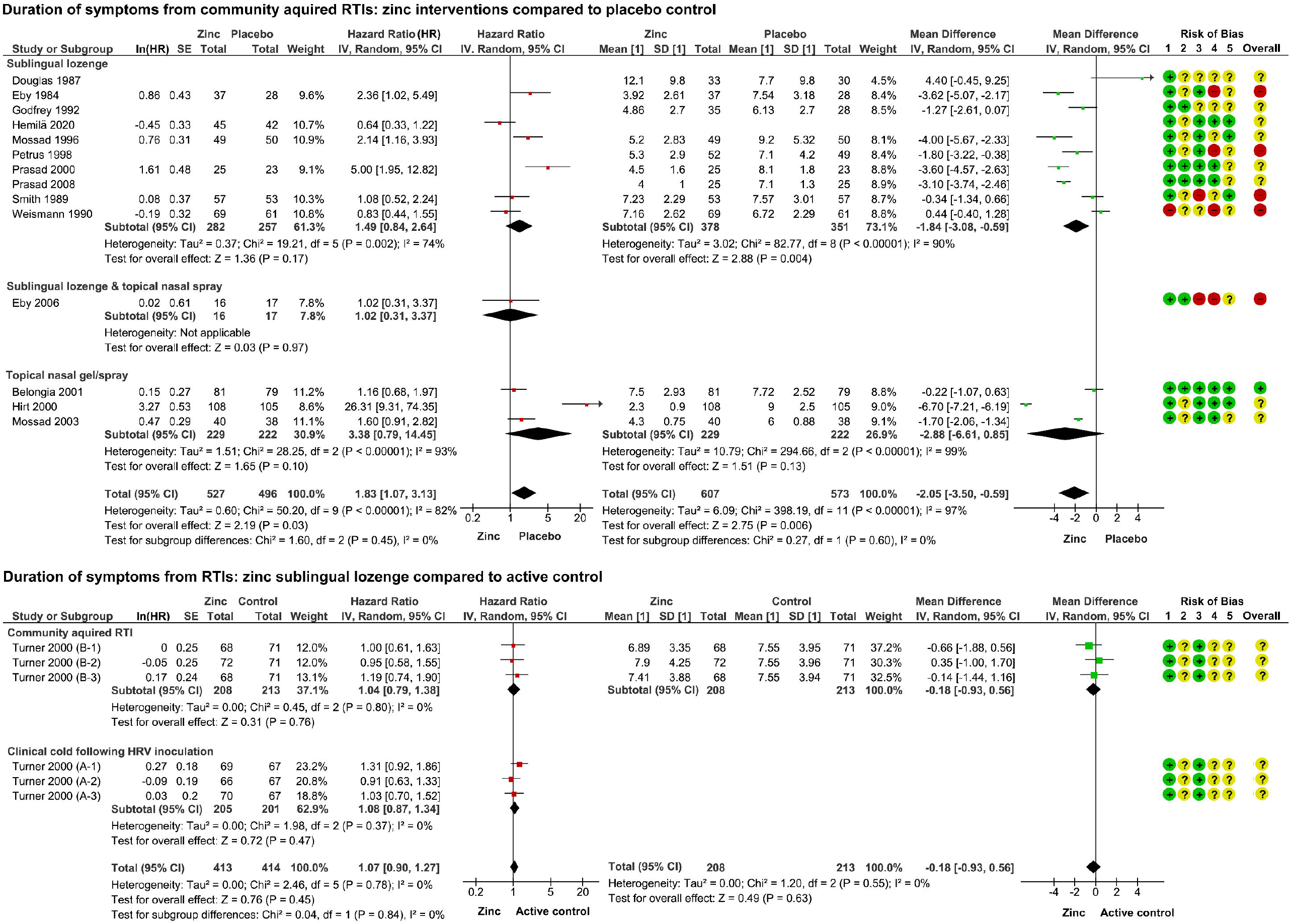
Duration of illness.

Results from the subgroup analyses that compared zinc salts, administration routes and zinc lozenge doses were less consistent. A significant difference between the zinc salt subgroups was found for mean duration (P=0.009) but not symptomatic risk (P=0.71). Similarly, a significant difference in the subgroups according to daily zinc lozenge dose was found for mean duration (P=0.002) but not symptomatic risk (P=0.27). No significant difference was found between the administration route subgroups for either mean duration (P=0.60) or symptomatic risk (P=0.27). To further investigate possible methodological explanations for the substantial heterogeneity, subgroup analyses were also conducted according to the maximum number of days participants were symptomatic prior to enrolment and different definitions of symptomatic survival, however, heterogeneity remained a concern.

The pooled results from two multicentre 4-arm RCTs that included 551 participants, found zinc lozenges were equivalent to an active control containing quinine (Figure 5). No differences were found in the likelihood of remaining symptomatic (HR 1.07, 95% CI 0.90 to 1.27, very low certainty SARS-CoV-2, moderate certainty other RTIs) from mild to moderately severe RTIs that were either community acquired or caused by HRV inoculation nor the mean days duration of illness for community acquired RTIs (MD −0.18 days, −0.93 to 0.56, very low certainty SARS-CoV-2, low certainty for other RTIs).

#### Adverse effects

No serious adverse effects were reported in any of the RCTs that evaluated zinc for treating RTIs (very low certainty SARS-CoV-2, moderate certainty for other RTIs).^67 70 73-86 92^ The risks of non-serious adverse effects were 1.4 times more for zinc compared to placebo (RR 1.41, 95% CI 1.17 to 1.69, very low certainty SARS-CoV-2, moderate certainty for other RTIs) in 11 RCTs including 1102 participants.^70 73 74 76 77 80^ ^84- 86^ (Figure 6).The pooled results of three RCTs that included 979 participants that compared various zinc interventions and doses with active controls, found no difference in the rates of non-serious adverse effects (RR 1.16, 95% CI 0.88 to 1.53).^67 92^

**FIGURE 6.**
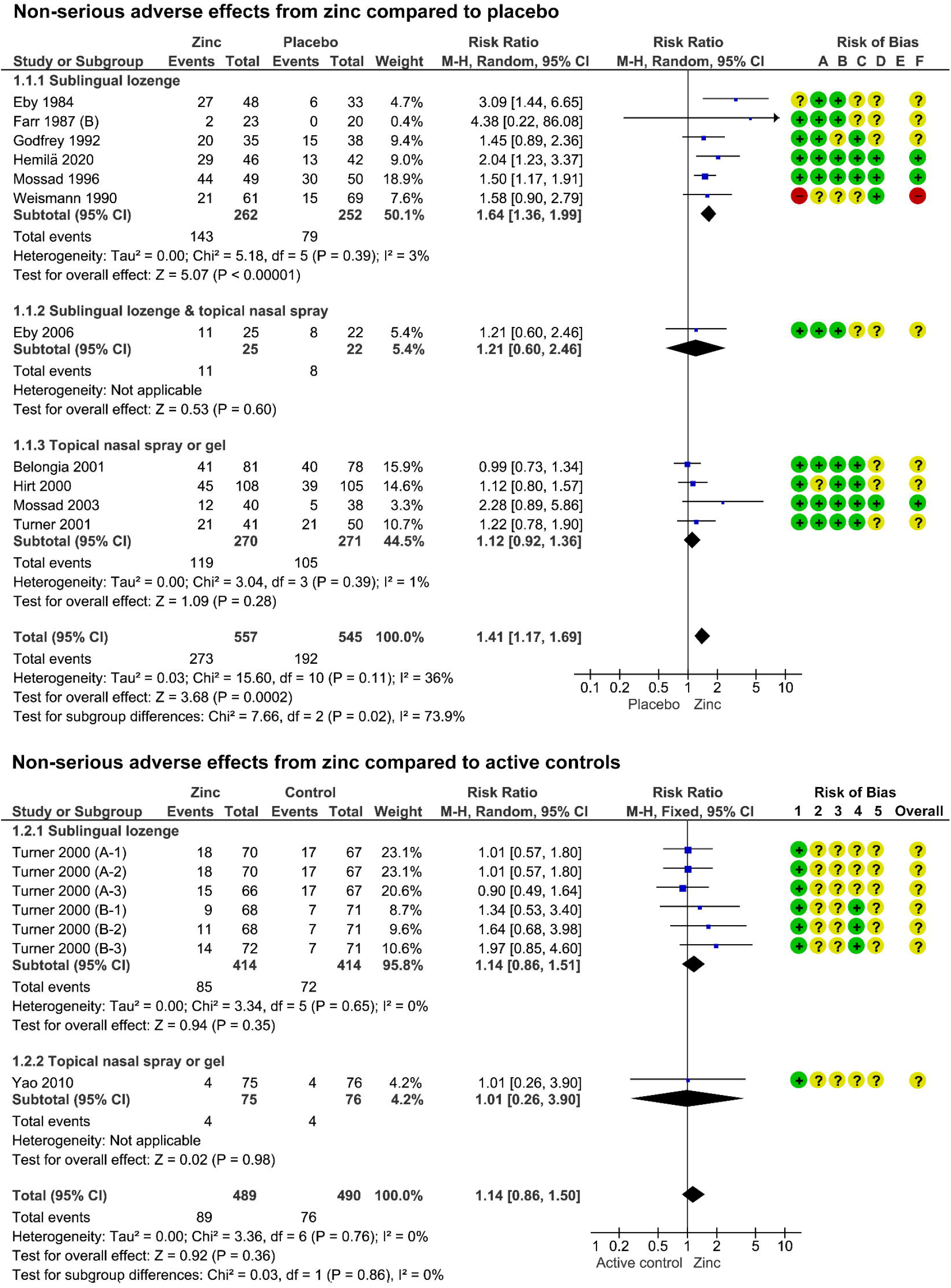
Adverse effects from zinc used to treat RTIs.

Sixteen RCTs reported the incidence of individual adverse effect rates. Nausea or gastro-intestinal discomfort was 1.3 times more likely with zinc compared to placebo (RR 1.30, 95% CI 1.01 to 1.90) found in 11 RCTs that included 957 participants.^70 73 75 77 79-81 84-87^ Mouth irritation or soreness from sublingual lozenges containing zinc, was 1.6 times more likely than placebo lozenges (RR 1.55, 95% CI 1.05 to 2.29) in seven RCTs that included 505 participants.^73 75 77 79 80 85 86^ Sublingual lozenges were more than twice as likely to cause taste aversion (RR 2.11, 95% CI 1.47 to 3.04) compared to placebo lozenges, in nine RCTs that included 719 participants.^73 75 77 79 80 82 84 85 87^ Zinc lozenges were more likely than nasal sprays and gels to cause any non-serious adverse effect (P=0.02) and/or gastrointestinal symptoms (P=0.04). Adverse effect incidence was inconsistent for comparisons of different zinc doses and salts compared to placebo controls. Unlike the prevention studies, the pooled results of three RCTs (328 participants) investigating zinc as a treatment,^74 76 83^ found no difference in the rates of nasal irritation or pain from zinc topical nasal sprays or gels compared to placebo controls (RR 1.22, 95% CI 0.72 to 2.05).

## DISCUSSION

### Principle findings

This rapid review and meta-analysis considers the potential role of zinc in the prevention and treatment of SARS-CoV-2, and advances the evidence on the effects of zinc for mild and moderate RTIs in adults without zinc deficiencies.^41-47^ New evidence about the prophylactic effects of zinc found that compared to placebo controls, zinc reduced the risk of developing symptoms that are consistent with a non-specific viral RTI. This risk was substantially lower for moderately severe illness compared to mild illness. When zinc was used to treat mild to moderate RTIs, there were clinically important reductions in the symptom severity and duration, however, the higher rates of non-serious adverse effects from zinc may limit its acceptability for some people.

Confidence in the estimated effects of zinc for SARS-CoV-2 is very low, due to serious indirectness as there were no data specific to SARS-CoV-2, nor other coronaviruses, and some concerns about bias for most outcomes. From the perspective of the general population, the indirect evidence is still relevant. The risk of other community acquired viral RTIs during the covid-19 pandemic remains relevant, and zinc is likely to be self-prescribed either before or near the onset of symptoms prior to knowing the cause.

Alongside potential benefits, consumers also seek detailed information about adverse effects and tolerability. The review confirms that adverse stomach, mouth, and nasal symptoms are common, particularly with doses above the NOEL of 40 to 50mg daily. No serious adverse effects were reported, suggesting this risk is low. However, post marketing surveillance has identified cases of long-lasting anosmia associated with a zinc gluconate nasal gel.^98 99^ It is unclear if the same risks apply to nasal sprays. A loss of smell was not reported by any of the RCTs, including the 1,364 young adults who used a zinc gluconate nasal spray twice daily for one month. As anosmia is an early SARS-CoV-2 symptom, any use of topical nasal zinc formulations during the pandemic should be considered and carefully monitored.^100^

Copper depletion is another potential risk, particularly with long-term zinc use,^29 30^ as might be the case for people choosing to use zinc for SARS-CoV-2 prophylaxis. This risk is reflected in the recommended upper limits of around 40 to 50mg daily.^30^ It was reassuring that plasma copper levels were stable following 15mg of oral zinc for seven months^88^ and 45mg of oral zinc daily for 12 months.^89^ However, both RCTs were small and may be underpowered to detect a difference, only a single marker of copper status was measured,^29^ and intestinal absorption of zinc is influenced by a variety of factors including age, medication use, dietary phytates (found in whole grains, legumes, nuts and seeds) and zinc status.^5 32^

### Implications for SARS-CoV-2 research

The review findings align with calls for more immuno-nutrition research, particularly in populations with a higher SARS-CoV-2 risk ^32 35^ Results from seven RCTs evaluating various zinc doses, salts, and administration routes for the prevention or treatment of SARS-CoV-2 are all pending. These RCTs will continue to be tracked and the review periodically updated until there is moderate certainty in the evidence or no further direct evidence is pending.

Future SARS-CoV-2 clinical trials should consider replicating the RCTs with positive results for other viral RTIs and consider focusing on high risk groups. Based on the limited information reported in the protocols, some of the choices for zinc interventions appear to be arbitrary. For instance, despite sublingual and topical nasal zinc demonstrating effectiveness for other viral RTIs, none of the registered SARS-CoV-2 trials plan to evaluate either administration route, and a relatively low dose of oral zinc, 15mg daily for prevention and 20mg daily for treatment, will be evaluated in two other RCTs despite neither dose demonstrating efficacy for other viral RTIs. Future studies could consider evaluating higher doses of prophylactic oral zinc (45mg daily in a divided dose) for older adults and people living in residential care, and low dose prophylactic zinc nasal spray for at risk younger adults, such as healthcare workers and contacts of known SARS-CoV-2 cases. For short-term, acute treatment of SARS-CoV-2, it would be reasonable to first investigate an oral or sublingual dose of at least 50mg, or perhaps even 75mg daily,^42 43^ or include multiple zinc arms with different doses. Regarding the choice of zinc salts to investigate, the largest body of evidence from other viral RTIs comes from zinc gluconate and zinc acetate, suggesting these are both suitable choices. Trialists should also ensure they classify zinc an active control and not an inert control.

The minimum time-frame in which zinc should be started following an acute SARS-CoV-2 infection will also need to be investigated. Most of the RCTs included in this review commenced zinc within 24 hours from the onset of symptoms. Yet, in the post-hoc subgroup analyses we conducted to evaluate methodological diversity the duration of illness was also reduced in the subset of RCTs in which participants commenced zinc up to three days from the onset of symptoms. Further, in a preliminary analysis for one of the included RCTs, the investigators briefly report that the significant reduction in the duration of symptoms remained when a large number of participants with symptoms of up to 10 days duration were added.^77^ Similarly, in an case series of four consecutive adults with probable or laboratory confirmed SARS-CoV-2 infection who took high dose zinc lozenges (115 – 207mg daily) as outpatients, only one person commenced zinc within 24 hours from the onset of symptoms. The other three commenced zinc five, nine and 21 days later. Noticeable clinical improvements over the next 24 hours were observed in all four cases.^20^ These findings suggest zinc could be beneficial even when it is commenced later in the course of an illness. SARS-CoV-2 studies will need to investigate if the duration of symptoms prior to commencing zinc impacts effectiveness.

Clues regarding timing and dose of zinc can be gleaned from a retrospective, open-label observational study of adults hospitalised with SARS-CoV-2.^17^ The study observed that a daily zinc dose of 100mg (zinc sulphate capsules, 220⍰mg twice daily for 5⍰days), as an adjuvant to hydroxychloroquine and azithromycin was associated with a decrease in mortality and hospice care. This association was strongest for adults who were not admitted to the intensive care unit (ICU). Since patients in ICU are experiencing more severe cytokine storms, the investigators postulate that zinc’s mechanism of action may be predominantly antiviral rather than anti-inflammatory. However, neither the duration of symptoms nor baseline zinc status were reported, nor were they included as explanatory variables. It should be considered that critically ill adults may have had a lower zinc status or required a higher zinc dose or perhaps a different administration route (e.g. intravenous).

Along with investigating the potential effectiveness of various doses, formulations and administration options, SARS-CoV-2 clinical trials should also determine if zinc requires a carrier or an ionophore, such as hydroxychloroquine,^16^ and compare the risks and benefits. According to our review findings and preliminary in vitro SARS-CoV-2 research,^33^ it is plausible that zinc may be effective when used on its own.

Finally, except for one RCT that evaluated the effects of zinc on cognitive function,^72 93^ the symptomatic and functional impact on the participants’ quality of life (QoL) was not assessed. Given the importance of these outcomes to patients, patient-reported outcomes measures such as the Wisconsin Upper Respiratory Symptom Survey (WURSS-24) that assess both symptom severity and quality of life are recommended.^101^

### Strengths and limitations of this review

Consultation with consumer advocates helped focus this review on issues that matter to patients and affirmed the relevance of indirect evidence. Notwithstanding, the indirectness to SARS-CoV-2 is a major limitation that constrained the overall certainty of the evidence. The only direct evidence that we are aware of comes from non-randomised studies of interventions.^8 16-19^ However, including these studied would not have changed the conclusions. The certainty of the evidence would have remained very low for all outcomes.

Other limitations to the certainty of the evidence included serious concerns about the inconsistency of the estimates of effect for duration. Wide variations in the methodology, zinc interventions, demographics of the study participants, and changes in the seasonal epidemiology of viral RTIs are all likely to have contributed to this heterogeneity. We were more cautious than previous reviewers,^45 46^ and downgraded the certainty of the evidence by two levels for inconsistency. A high risk of bias for most of the RCT outcomes was another concern. Except for adverse effects, many RCT outcomes has at least some concerns. Reassuringly, the estimates of effects for all critical outcomes were robust following removal of RCTs with a high risk of bias. Nevertheless, were more cautious than previous reviewers,^42-46^ and downgraded the certainty of the evidence due to risk of bias by one level for all prevention, duration and severity outcomes.

Limitations of the rapid review methods included single reviewers conducting many of the tasks. This may have increased errors and inconsistencies. Strategies to minimise these risks included calibrating the reviewers prior to conducting single tasks, applying a high threshold for exclusion when screening articles, and a second reviewer verifying data extraction and appraisal. Other errors may have arisen from using second-hand information reported in other systematic reviews to augment the analysis of non-critical outcomes and subgroup analyses.^41-43^ As this review is the first to analyse hazard ratios, including this data meant the pooled results could be compared to the alternate effect measure, mean days duration.

Rapid review limitations were counterbalanced by not restricting the search strategy and including adults with viral RTIs following HRV inoculation that led to substantially more studies being included compared to previous reviews.^41-47^ Notably, a concurrent systematic review published in 2020 only included 1 of the 6 prevention RCTs and 9 of the 23 treatment RCTs.^46^ This in part was due to the reviewers only searching four databases and affirms the importance of carefully considering which review methods to restrict.^102^

The decision to use the RoB-2 tool increased the workload, as RCTs with multiple outcomes were appraised more than once. Hence, only the outcomes included in a meta-analysis were appraised. The trade-off of appraising bias at the outcome level rather than study level was the improved specificity and relevance of RoB-2 to zinc interventions. For example, missing responses to questionnaire items were more likely to bias symptoms severity score than symptom duration. The signalling questions for Domain 2: Deviations from Intended Interventions, helped highlight bias arising from contamination of the intervention, a notable challenge in the evaluation of nutritional (dietary) interventions.

Flexibility in the rapid review protocol enabled a staged meta-analysis and more timely reporting of priority conditions, populations, and outcomes. The 95 shortlisted RCTs evaluating zinc for the prevention or treatment of viral RTIs in children are yet to be analysed, and will add to the findings of the 2016 Cochrane review that included six RCTs evaluating zinc supplementation for the prevention of pneumonia in children aged from 2 months to 59 months.^48^ In contrast to the RCTs of adult populations, most RCTs evaluated zinc use in populations with a moderate or high risk of zinc deficiency, and around one third were prevention studies.

## CONCLUSIONS

This rapid review of RCTs confirms that zinc is a potential therapeutic candidate for preventing and treating SARS-CoV-2, including high risk groups and adults without zinc deficiency. The seven trials currently in progress, will provide important direct evidence for SARS-CoV-2. Pending these results, the indirect evidence from this review points towards zinc being a viable alternative to other unproven pharmaceutical options. Trials comparing standard care to zinc alone and as an adjuvant to other interventions are both warranted. The choice of zinc interventions to investigate should be guided by the findings of this review. Along with clinical outcomes, quality of life and adverse effects should be assessed. Additional information about the relative availability, acceptability and costs of therapeutic options will also be needed.

## Data Availability

Additional data is available upon reasonable request to the authors

